# Microbiological characteristics of ventilator-associated pneumonia in patients with neurological injuries: a multicentric cohort

**DOI:** 10.64898/2026.10.15.26365620

**Authors:** Lucie Foueré, Maëlle Youinou, Caroline Schimpf, Nicolas Mongardon, Christophe Quesnel, François Philippart

**Affiliations:** Medical and surgical intensive care department. Groupe hospitalier Paris Saint Joseph. Paris, France; Department of Clinical Research, Groupe Hospitalier Paris Saint Joseph, Paris, France; Anaesthesiology and intensive care Department, Sainte Anne Hospital, Paris, France; Department of Anesthesiology, Surgical Intensive Care, and perioperative Medicine, Henri Mondor hospital, Créteil, France; INSERM, Pharmacologie et Technologies pour les Maladies Cardiovasculaires (PROTECT), École Nationale Vétérinaire d’Alfort (EnvA), Université Paris Est Créteil (UPEC)’’, Créteil, 94010, France; Faculty of Health Sciences, University of Paris Est Créteil, Créteil, F-94010, France; GRCT OPTIMA, Université Paris Est Créteil, Créteil, F-94010, France; Endotoxins, Structures and Host Response, Department of Microbiology, Institute for Integrative Biology of the Cell, UMR 9891 CNRS-CEA-Paris Saclay University, 98190 Gif-sur-Yvette, France

**Keywords:** *Staphylococcus aureus*, Ventilator-associated pneumonia, brain injury, ICU

## Abstract

Incidence of ventilator-associated pneumonia (VAP) is increased in patients admitted for brain injury. *Staphylococcus aureus* appears to be a particularly common pathogen in these respiratory infections. Our aim was to confirm the extent of *S. aureus* involvement in VAP among patients with neurological injuries. We conducted a retrospective, multicenter study including patients with and without neurological injuries who developed VAP during their ICU stay. Among 15 702 patients admitted in the participant centers during the study period. 407 were finally included in the study. *S. aureus* was involved in 122 (61.6%) VAP in brain-injured patients and 63 (30.1%) of non-brain-injured patients. In brain-injured patients, multivariate analysis identified age (0.97 [0.95; 0.98]), antibioprophylaxis (0.31 [0.18; 0.53]; p < 0.0001), subarachnoid hemorrhage (2.18 [1.01; 4.68]; p=0.0461), lower Glasgow coma score (0.92 [0.87; 0.97]; p= 0.004) and SAPS II (0.98 [0.97; 1]; p=0.0174) as critical factors associated with the risk of *S. aureus* in VAP. In the non-brain-injured population, multivariate analysis identified antibioprophylaxis (0.38 [0.22; 0.65]; p=0.0006), age (0.96 [0.95; 0.98]; p<0.0001), SAPS II (0.98 [0.97; 1]; p=0.0265), thoracic surgery (0.21 [0.09; 0.48]; p=0.0002) and peripheral neurosurgery (1.95 [1.2; 3.19]; p=0.0076) as critical factors. A total of 172 patients (42.3%) died during their ICU stay and 90-days mortality was 194 (47.7%). No difference of mortality was noticed function of VAP-associated pathogen.

## Introduction

Despite structured bundle of prevention^1,2^, ventilator-associated pneumonia remains the most common nosocomial infection in intensive care units^3,4^ and they account for half of the antibiotherapy administrered in ICU^5^. VAP is associated with an increase of mechanical ventilation duration^6–8^ and ICU length of stay^6,8–10^. Although attributable mortality appears modest^8,11–13^, the crude mortality rate associated with VAP remains significant^9,10^. It is even higher when initial antibiotic therapy has been ineffective^14–19^.

In patients with brain injuries, the risk of developing VAP is up to 12 times higher than in the general intensive care population^20^. The incidence of VAP in this setting ranges from 20 to more than 50%^20–26^, or 7 to more than 33 per 1,000 days of mechanical ventilation^24,27^. The incidence varies depending on the type of injury, being higher for traumatic brain injury and ischemic strokes (30 to 60%)^20,22,24,26^, hemorrhagic strokes (around 30%)^24,27^ than for subarachnoid hemorrhages (approximately 20%)^24^. Numerous factors contribute to the development of infection, particularly pneumonia, in patients with neurological injuries^24,25^. These factors include an impairment of the immune response and its regulation^28,29^— summarized by the concept of brain-induced immunodepression^30–33^— as well as alterations in the visceral brain-lung^34^, brain-gut^35^, and gut-lung ^36^axes, changes in the microbiomes^37–39^, and the risks of translocation and inhalation^40^. These infections are associated with an increase in the duration of mechanical ventilation and sedation, the need for tracheostomy^20,21,23,24^, in-hospital mortality^20^; and a poor long-term prognosis^20,26,41^ favored by cerebral oxygenation compromission^26^.

In addition to their particularly high prevalence in cases of brain injury, VAP appear to have specific microbiological characteristics. *Staphylococcus aureus* is a major pathogen in early-onset VAP^9,42,43^. Its role appears to be particularly significant in brain injured patients, during early-onset sepsis^44^ and more specifically during VAP. and may account for more than 40% of the pathogens identified^20,21,26,45–47^.

Our objective is to confirm the significance of *S. aureus* involvement among the pathogens responsible for early-onset VAP in the neuroinjured population and to identify the risk factors associated with its involvement in VAP among neuroinjured patients.

## Method

### Study settings

The study adhered to the STROBE (STrengthening the Reporting of Observational Studies in Epidemiology) guidelines for reporting observational studies. We conduct a retrospective multicentric observational study in three critical care departments of three regional university hospital in Paris.

### Patients selection / Participants

Between January 1^st^ 2021 and June 1^st^ 2025 all consecutive adult patients admitted in one of the participant departments and who required an invasive mechanical ventilation of at least 48h were screened for inclusion.

### Endpoints

The primary objective of the study was to confirm the predominance of *S. aureus* among VAP occurring in brain-injured patients and to identify the factors associated with the risk of *S. aureus* involvement during VAP in this context. Risk factors for involvement of *S. aureus* in non-brain-injured patients associated VAP; ICU and 90 days mortality associated with involvement of S. aureus, and the association of association with other pathogens, were also investigated.

### Definitions

The same definitions were used during the whole period of the study. Hospital acquired and VAP were defined according to the European^48,49^ and American guidelines^50^. Microbiological confirmation was obtained by sputum culture (≥10^7^ cfu/ml), tracheal aspirate (≥10^6^ cfu/ml), or, in ventilated patients, bronchoalveolar lavage (≥10^4^ cfu/ml)^50–52^.

### Microbiology

All bacterial isolates were sent to the corresponding hospital’s microbiology laboratory. Bacterial identification was performed using the MALDI-TOF mass spectrometry (Andromas system, Paris, France). Antimicrobial susceptibility testing (AST) was performed according to the CA-SFM guidelines (*Comité de l’Antibiogramme de la Société Française de Microbiologie*) (2011-2014) or European Committee on Antimicrobial Susceptibility Testing – EUCAST– (2015-2016) by disk diffusion method with Mueller Hinton agar (Biorad, Marnes-la–Coquette, France), using McFarland 0.5 from overnight cultures, followed by incubation at 35°C for 16 to 18 h. Diameters were interpreted accordingly to the breakpoints of CA-SFM or EUCAST guidelines.

Classification of *Enterobacteriaceae* used in the tables refers the natural resistance to beta-lactamins^53,54^.

### Data collection

Data were collected from electronic medical files, using a standardized electronic form. The severity of neurological conditions is assessed using the severity scores commonly employed in relevant intensive care units. The Glasgow Coma Scale^55^ was recorded in all cases. The severity of ischemic strokes was assessed using the National Institutes of Health Stroke Scale (NIHSS)^56^; that of subarachnoid hemorrhages, using the Fisher score^57^ and the World Federation of Neurosurgical Societies (WFNS) scale^58^; and that of **traumatic brain injuries, using**. The demographic data, the reason for invasive ventilation, and the clinical and laboratory parameters, as well as the therapeutic interventions collected, are detailed in the supplementary material.

### Ethics

The study was carried out in accordance with the Declaration of Helsinki and applicable French law. It was approved by our institutional review board (GERM—*Groupe Ethique & Recherche Médicale*—; IRB n°00 012 157). The requirement for written informed consent was waived by the GERM. Participants Patients were informed about the study and their non-opposition to the collection and use of non-identifying data for research purposes was obtained. Data processing complied with the reference methodology MR-004 of the French *Commission Nationale de l’Informatique et des Libertés* (French National Data Protection Agency).

### Study size

Based on the literature and our preliminary data, the risk of developing a VAP caused by Gram-positive cocci was approximately 25% in intensive care patients without brain injury, and approximately 40% in the population with brain injury. We estimated that to detect a difference in the presence of *S. aureus* in VAP cases based on the presence or absence of brain injury, with a 5% alpha level and 90% power using a two-sided chi-square test, a sample size of approximately 400 patients would be required. Furthermore, this population was also intended to identify potential risk factors for S. aureus involvement in these PAVMs during multivariate analysis.

### Statistical analysis

Quantitative variables were presented as means (+/− standard deviation) or medians [interquartile range] and minimum-maximum, depending on their distribution, evaluated using visual inspection and normality tests. Qualitative variables were described in terms of numbers and percentages. When appropriate, differences in means, medians, or proportions between groups were reported with 95% confidence intervals (95% CI), computed using normal approximation or bootstrap methods for means and medians, and the Clopper-Pearson method for proportions. Univariate analysis: A list of candidate risk factors was defined a priori. Odds ratios (ORs) with their 95% CI were computed on the original data, and p-values were calculated using the likelihood ratio test. Variables with a p-value < 0.20 were selected. No variables were forced into the model. Multivariate analysis: The final multivariate model was obtained using a backward stepwise selection procedure, applied independently to each of the 40 imputed datasets to account for imputation-related uncertainty during the variable selection process. A majority rule was then applied: only variables selected in at least 50% of the imputed datasets were retained in the final model. The resulting model was subsequently refit to each imputed dataset, and the coefficient estimates were pooled using Rubin’s rules to derive adjusted odds ratios and their 95% CI. Internal validation: Optimism correction was performed using a bootstrap procedure adapted to multiply imputed data (200 replications), with the complete modelling process (univariate screening, backward stepwise selection, model fitting, pooling using Rubin’s rules) repeated at each replication. Model performance was assessed both on the bootstrap and original imputed datasets, and optimism was estimated as the average difference between these two performances across replications. Discrimination was assessed using the c-statistic, and calibration using the calibration slope and the observed-to-expected (O/E) ratio; Cox and Snell’s R² was used as a measure of explained variation. Sensitivity analysis: A sensitivity analysis was performed using complete-case analysis, to assess the robustness of the ORs from the multiple imputation-based model. The significance threshold has been set at p < 0.05. Statistical analysis were performed using the R® software (R v4.4.2, URL: https://www.R-project.org/). Detail are provided in supplementary material.

## Results

### Participants

Between January first 2021 and June first 2025, a total of 15 702 patients were admitted in the participant centers during the study period. Among them 923 developed a VAP. A total of 516 were excluded (cf. **figure 1**). Among the remaining 407 patients, 198 were admitted in ICU with a brain injury and 209 for other reasons. The acute brain lesion was a trauma in 79 (39.9%%), an ischemic stroke in 20 (10.1%), a hematoma in 42 (21.2%) or a sub-arachnoid hemorrhage in 53 (26.8%). Details in **table 1**. The motif of admission of non-brain-injured patients were mainly acute respiratory failure in 85 (40.7%), polytrauma in 20 (9.6%) infection (sepsis or septic shock) in 10 (4.8%), and planned surgery in 49 (23.4%). Details in **table 1**.

**Table 1:**
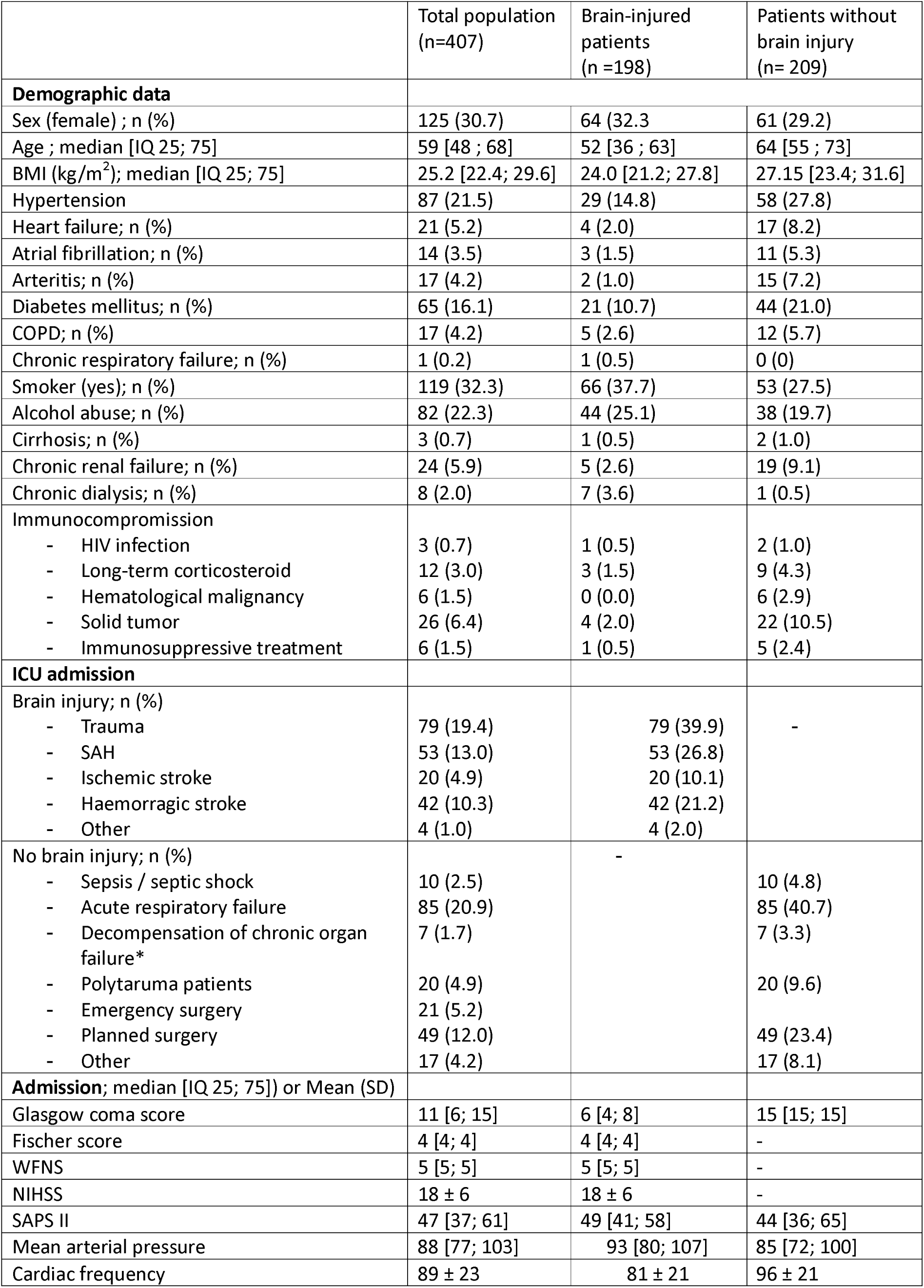
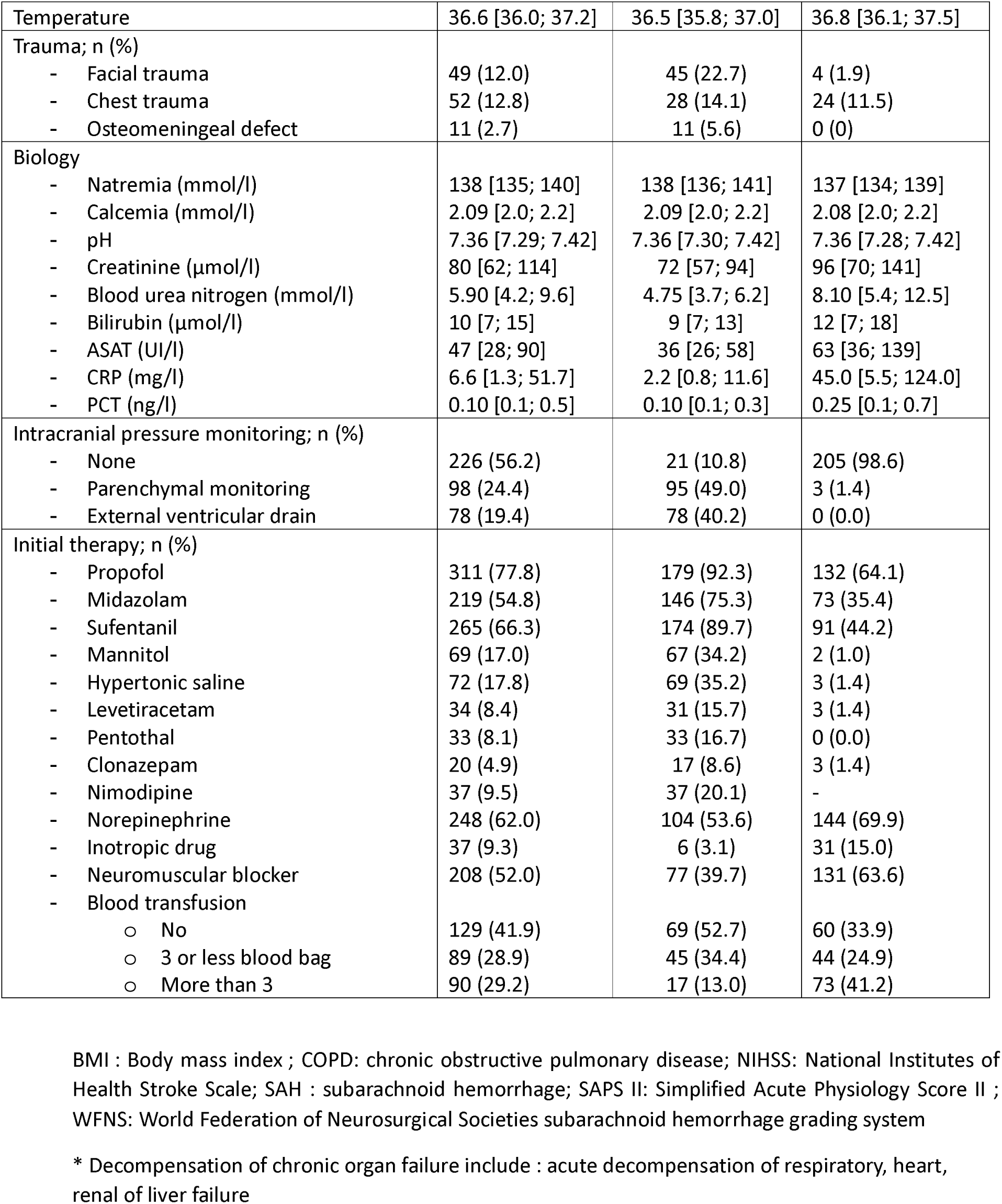
demographic characteristics.

|  | Total population<br>(n=407) | Brain-injured<br>patients<br>(n =198) | Patients without<br>brain injury<br>(n= 209) |
| --- | --- | --- | --- |
| **Demographic data** |  |  |  |
| Sex (female) ; n (%) | 125 (30.7) | 64 (32.3) | 61 (29.2) |
| Age ; median [IQ 25; 75] | 59 [48 ; 68] | 52 [36 ; 63] | 64 [55 ; 73] |
| BMI (kg/m<sup>2</sup>); median [IQ 25; 75] | 25.2 [22.4; 29.6] | 24.0 [21.2; 27.8] | 27.15 [23.4; 31.6] |
| Hypertension | 87 (21.5) | 29 (14.8) | 58 (27.8) |
| Heart failure; n (%) | 21 (5.2) | 4 (2.0) | 17 (8.2) |
| Atrial fibrillation; n (%) | 14 (3.5) | 3 (1.5) | 11 (5.3) |
| Arteritis; n (%) | 17 (4.2) | 2 (1.0) | 15 (7.2) |
| Diabetes mellitus; n (%) | 65 (16.1) | 21 (10.7) | 44 (21.0) |
| COPD; n (%) | 17 (4.2) | 5 (2.6) | 12 (5.7) |
| Chronic respiratory failure; n (%) | 1 (0.2) | 1 (0.5) | 0 (0) |
| Smoker (yes); n (%) | 119 (32.3) | 66 (37.7) | 53 (27.5) |
| Alcohol abuse; n (%) | 82 (22.3) | 44 (25.1) | 38 (19.7) |
| Cirrhosis; n (%) | 3 (0.7) | 1 (0.5) | 2 (1.0) |
| Chronic renal failure; n (%) | 24 (5.9) | 5 (2.6) | 19 (9.1) |
| Chronic dialysis; n (%) | 8 (2.0) | 7 (3.6) | 1 (0.5) |
| Immunocompromission |  |  |  |
| - HIV infection | 3 (0.7) | 1 (0.5) | 2 (1.0) |
| - Long-term corticosteroid | 12 (3.0) | 3 (1.5) | 9 (4.3) |
| - Hematological malignancy | 6 (1.5) | 0 (0.0) | 6 (2.9) |
| - Solid tumor | 26 (6.4) | 4 (2.0) | 22 (10.5) |
| - Immunosuppressive treatment | 6 (1.5) | 1 (0.5) | 5 (2.4) |
| **ICU admission** |  |  |  |
| Brain injury; n (%) |  |  |  |
| - Trauma | 79 (19.4) | 79 (39.9) | - |
| - SAH | 53 (13.0) | 53 (26.8) |  |
| - Ischemic stroke | 20 (4.9) | 20 (10.1) |  |
| - Haemorrhagic stroke | 42 (10.3) | 42 (21.2) |  |
| - Other | 4 (1.0) | 4 (2.0) |  |
| No brain injury; n (%) |  | - |  |
| - Sepsis / septic shock | 10 (2.5) |  | 10 (4.8) |
| - Acute respiratory failure | 85 (20.9) |  | 85 (40.7) |
| - Decompensation of chronic organ failure* | 7 (1.7) |  | 7 (3.3) |
| - Polytrauma patients | 20 (4.9) |  | 20 (9.6) |
| - Emergency surgery | 21 (5.2) |  |  |
| - Planned surgery | 49 (12.0) |  | 49 (23.4) |
| - Other | 17 (4.2) |  | 17 (8.1) |
| **Admission; median [IQ 25; 75]) or Mean (SD)** |  |  |  |
| Glasgow coma score | 11 [6; 15] | 6 [4; 8] | 15 [15; 15] |
| Fischer score | 4 [4; 4] | 4 [4; 4] | - |
| WFNS | 5 [5; 5] | 5 [5; 5] | - |
| NIHSS | 18 ± 6 | 18 ± 6 | - |
| SAPS II | 47 [37; 61] | 49 [41; 58] | 44 [36; 65] |
| Mean arterial pressure | 88 [77; 103] | 93 [80; 107] | 85 [72; 100] |
| Cardiac frequency | 89 ± 23 | 81 ± 21 | 96 ± 21 |
| Temperature | 36.6 [36.0; 37.2] | 36.5 [35.8; 37.0] | 36.8 [36.1; 37.5] |
| Trauma; n (%) |  |  |  |
| - Facial trauma | 49 (12.0) | 45 (22.7) | 4 (1.9) |
| - Chest trauma | 52 (12.8) | 28 (14.1) | 24 (11.5) |
| - Osteomeningeal defect | 11 (2.7) | 11 (5.6) | 0 (0) |
| Biology |  |  |  |
| - Natremia (mmol/l) | 138 [135; 140] | 138 [136; 141] | 137 [134; 139] |
| - Calcemia (mmol/l) | 2.09 [2.0; 2.2] | 2.09 [2.0; 2.2] | 2.08 [2.0; 2.2] |
| - pH | 7.36 [7.29; 7.42] | 7.36 [7.30; 7.42] | 7.36 [7.28; 7.42] |
| - Creatinine (μmol/l) | 80 [62; 114] | 72 [57; 94] | 96 [70; 141] |
| - Blood urea nitrogen (mmol/l) | 5.90 [4.2; 9.6] | 4.75 [3.7; 6.2] | 8.10 [5.4; 12.5] |
| - Bilirubin (μmol/l) | 10 [7; 15] | 9 [7; 13] | 12 [7; 18] |
| - ASAT (UI/l) | 47 [28; 90] | 36 [26; 58] | 63 [36; 139] |
| - CRP (mg/l) | 6.6 [1.3; 51.7] | 2.2 [0.8; 11.6] | 45.0 [5.5; 124.0] |
| - PCT (ng/l) | 0.10 [0.1; 0.5] | 0.10 [0.1; 0.3] | 0.25 [0.1; 0.7] |
| Intracranial pressure monitoring; n (%) |  |  |  |
| - None | 226 (56.2) | 21 (10.8) | 205 (98.6) |
| - Parenchymal monitoring | 98 (24.4) | 95 (49.0) | 3 (1.4) |
| - External ventricular drain | 78 (19.4) | 78 (40.2) | 0 (0.0) |
| Initial therapy; n (%) |  |  |  |
| - Propofol | 311 (77.8) | 179 (92.3) | 132 (64.1) |
| - Midazolam | 219 (54.8) | 146 (75.3) | 73 (35.4) |
| - Sufentanil | 265 (66.3) | 174 (89.7) | 91 (44.2) |
| - Mannitol | 69 (17.0) | 67 (34.2) | 2 (1.0) |
| - Hypertonic saline | 72 (17.8) | 69 (35.2) | 3 (1.4) |
| - Levetiracetam | 34 (8.4) | 31 (15.7) | 3 (1.4) |
| - Pentothal | 33 (8.1) | 33 (16.7) | 0 (0.0) |
| - Clonazepam | 20 (4.9) | 17 (8.6) | 3 (1.4) |
| - Nimodipine | 37 (9.5) | 37 (20.1) | - |
| - Norepinephrine | 248 (62.0) | 104 (53.6) | 144 (69.9) |
| - Inotropic drug | 37 (9.3) | 6 (3.1) | 31 (15.0) |
| - Neuromuscular blocker | 208 (52.0) | 77 (39.7) | 131 (63.6) |
| - Blood transfusion |  |  |  |
| o No | 129 (41.9) | 69 (52.7) | 60 (33.9) |
| o 3 or less blood bag | 89 (28.9) | 45 (34.4) | 44 (24.9) |
| o More than 3 | 90 (29.2) | 17 (13.0) | 73 (41.2) |

BMI : Body mass index ; COPD: chronic obstructive pulmonary disease; NIHSS: National Institutes of Health Stroke Scale; SAH : subarachnoid hemorrhage; SAPS II: Simplified Acute Physiology Score II ; WFNS: World Federation of Neurosurgical Societies subarachnoid hemorrhage grading system
\* Decomensation of chronic organ failure include : acute decompensation of respiratory, heart, renal of liver failure

In brain-injured patients, initial Glasgow coma score was 6 [4; 8]. Fischer score of sub-arachnoid hemorrhage was 4 [4; 4], WFNS was 5 [5; 5], and NIHSS score in strokes was 18 ± 6. Details in **table 1**. Specificities of care function of type of brain injury are detailed in supplementary table 1.

Initial interventions required mechanical ventilation in the whole population. Norepinephrine in 208 (52.0%), inotropic drug in 37 (9.3%), blood transfusion in 278 (59.1%) and neuromuscular blocker in 208 (52.0). Specifically, in brain-injured subpopulation, mannitol was required in 67 (34.2%), hypertonic saline in 69 (35.2%), nimodipine in 37 (20.1, levetiracetam in 31 (15.7%) and Pentothal in 33 (16.7). Details il **table 1**.

### Ventilator-associated pneumonia

All patients developed a VAP during ICU stay. The delay to VAP onset was 6 [4; 9] in the whole population, 6 [4; 8] among brain-injured patients and 6 [4; 9] in the patient without brain insult.

At VAP onset an ARDS was present in 175 (43.0%) of patients (51 (25.8%) of brain-injured and 124 (59.3%) without brain injury), a septic shock in 174 (44.1%) (32.0% in brain-injured vs 55.7% without brain injury). Patients received previous antibiotic therapy by beta-lactamins in 151 (37.3%) (17.3% in brain-injured vs 56.0% without brain injury). An enteral nutrition was present in 273 (98.2%) of patients; systemic corticosteroids were present in 164 (40.3%) (26.7% *vs* 53.1%) and proton pump inhibitors in 97 (23.8%) (16.2% *vs* 31.1%). Details in **table 2**.

**Table 2:**
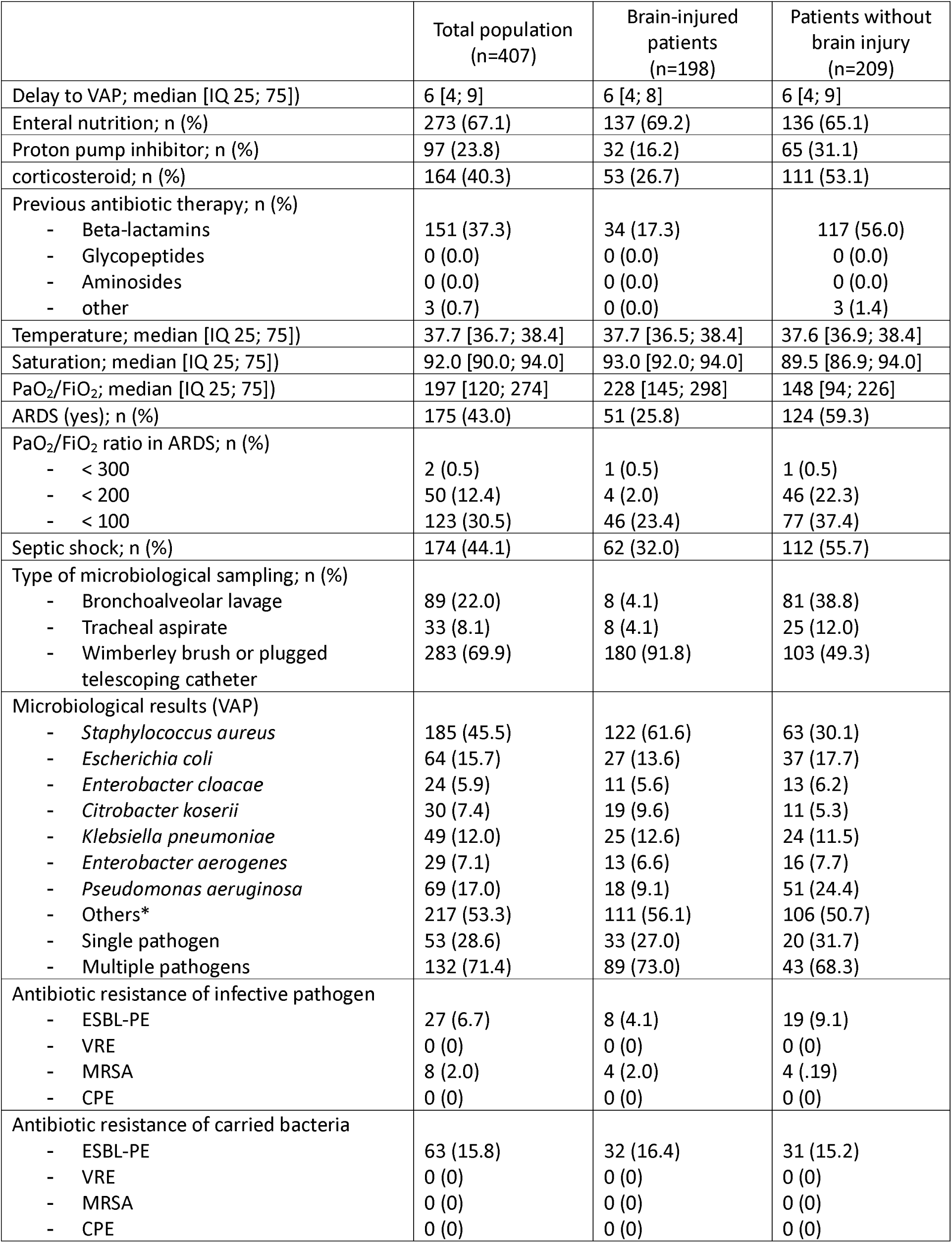
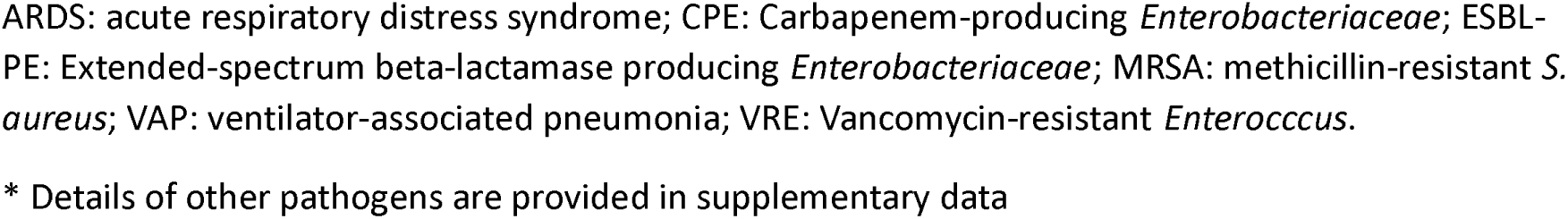
population at the time of VAP.

|  | Total population<br>(n=407) | Brain-injured<br>patients<br>(n=198) | Patients without<br>brain injury<br>(n=209) |
| --- | --- | --- | --- |
| Delay to VAP; median [IQ 25; 75]) | 6 [4; 9] | 6 [4; 8] | 6 [4; 9] |
| Enteral nutrition; n (%) | 273 (67.1) | 137 (69.2) | 136 (65.1) |
| Proton pump inhibitor; n (%) | 97 (23.8) | 32 (16.2) | 65 (31.1) |
| corticosteroid; n (%) | 164 (40.3) | 53 (26.7) | 111 (53.1) |
| Previous antibiotic therapy; n (%) |  |  |  |
| - Beta-lactamins | 151 (37.3) | 34 (17.3) | 117 (56.0) |
| - Glycopeptides | 0 (0.0) | 0 (0.0) | 0 (0.0) |
| - Aminoglycosides | 0 (0.0) | 0 (0.0) | 0 (0.0) |
| - other | 3 (0.7) | 0 (0.0) | 3 (1.4) |
| Temperature; median [IQ 25; 75]) | 37.7 [36.7; 38.4] | 37.7 [36.5; 38.4] | 37.6 [36.9; 38.4] |
| Saturation; median [IQ 25; 75]) | 92.0 [90.0; 94.0] | 93.0 [92.0; 94.0] | 89.5 [86.9; 94.0] |
| PaO<sub>2</sub>/FiO<sub>2</sub>; median [IQ 25; 75]) | 197 [120; 274] | 228 [145; 298] | 148 [94; 226] |
| ARDS (yes); n (%) | 175 (43.0) | 51 (25.8) | 124 (59.3) |
| PaO<sub>2</sub>/FiO<sub>2</sub> ratio in ARDS; n (%) |  |  |  |
| - < 300 | 2 (0.5) | 1 (0.5) | 1 (0.5) |
| - < 200 | 50 (12.4) | 4 (2.0) | 46 (22.3) |
| - < 100 | 123 (30.5) | 46 (23.4) | 77 (37.4) |
| Septic shock; n (%) | 174 (44.1) | 62 (32.0) | 112 (55.7) |
| Type of microbiological sampling; n (%) |  |  |  |
| - Bronchoalveolar lavage | 89 (22.0) | 8 (4.1) | 81 (38.8) |
| - Tracheal aspirate | 33 (8.1) | 8 (4.1) | 25 (12.0) |
| - Wimberley brush or plugged telescoping catheter | 283 (69.9) | 180 (91.8) | 103 (49.3) |
| Microbiological results (VAP) |  |  |  |
| - *Staphylococcus aureus* | 185 (45.5) | 122 (61.6) | 63 (30.1) |
| - *Escherichia coli* | 64 (15.7) | 27 (13.6) | 37 (17.7) |
| - *Enterobacter cloacae* | 24 (5.9) | 11 (5.6) | 13 (6.2) |
| - *Citrobacter koserii* | 30 (7.4) | 19 (9.6) | 11 (5.3) |
| - *Klebsiella pneumoniae* | 49 (12.0) | 25 (12.6) | 24 (11.5) |
| - *Enterobacter aerogenes* | 29 (7.1) | 13 (6.6) | 16 (7.7) |
| - *Pseudomonas aeruginosa* | 69 (17.0) | 18 (9.1) | 51 (24.4) |
| - Others* | 217 (53.3) | 111 (56.1) | 106 (50.7) |
| - Single pathogen | 53 (28.6) | 33 (27.0) | 20 (31.7) |
| - Multiple pathogens | 132 (71.4) | 89 (73.0) | 43 (68.3) |
| Antibiotic resistance of infective pathogen |  |  |  |
| - ESBL-PE | 27 (6.7) | 8 (4.1) | 19 (9.1) |
| - VRE | 0 (0) | 0 (0) | 0 (0) |
| - MRSA | 8 (2.0) | 4 (2.0) | 4 (1.9) |
| - CPE | 0 (0) | 0 (0) | 0 (0) |
| Antibiotic resistance of carried bacteria |  |  |  |
| - ESBL-PE | 63 (15.8) | 32 (16.4) | 31 (15.2) |
| - VRE | 0 (0) | 0 (0) | 0 (0) |
| - MRSA | 0 (0) | 0 (0) | 0 (0) |
| - CPE | 0 (0) | 0 (0) | 0 (0) |

ARDS: acute respiratory distress syndrome; CPE: Carbapenem-producing *Enterobacteriaceae*; ESBL-PE: Extended-spectrum beta-lactamase producing *Enterobacteriaceae*; MRSA: methicillin-resistant *S. aureus*; VAP: ventilator-associated pneumonia; VRE: Vancomycin-resistant *Enterococcus*.
\* Details of other pathogens are provided in supplementary data

Microbiological sample was obtained by bronchoalveolar lavage in 89 (22.0%) of cases (4.1% in the brain-injured population and 38.8% in the non-brain-injured patients), Wimberley brush or plugged telescoping catheter in 283 (69.9%) (91.8% vs 49.3%), or tracheal aspirate in 33 (8.1%) (4.1% vs 12.0%). Details in **table 2**.

The most commonly associated pathogen in our population were *Staphylococcus aureus* in 185 (45.5%), *Pseudomonas aeruginosa* in 69 (17.0%), *Escherichia coli* in 64 (15.7%) and *Klebsiella pneumoniae* in 49 (12.0%). Pathogens involved in the VAP episode were detailed in **table 2**. Among VAP involving *S. aureus* (n=185), a single pathogen was found in 53 (28.6%) whereas multiples pathogens were identified in 132 (71.4%). The most frequent pathogen associated were *E. coli* (24; 18.2%), *K. pneumoniae* (21; 15.9%), *Enterobacter aerogenes* (15; 11.4%) and *P. aeruginosa* (11; 8.3%). Details are provided in **table 3** and **supplementary table 2**. Repartition of responsible pathogens function of type of brain injury are available in **supplementary table 3**.

**Table 3:** Associated pathogen during S. aureus VAP.

| Associated pathogen | n (%) | Time to VAP<br>IQ [25;75] |
| --- | --- | --- |
| *E. coli* | 24 (18.2) | 6.0 [4.0; 8.0] |
| *E. cloacae* | 9 (6.8) | 7.0 [4.0; 8.0] |
| *C. koserii* | 12 (9.1) | 6.5 [4.5; 10.0] |
| *K. pneumoniae* | 21 (15.9) | 6.0 [4.0; 10.0] |
| *E. aerogenes* | 15 (11.4) | 6.0 [5.0; 9.0] |
| *P. aeruginosa* | 11 (8.3) | 5.0 [3.0; 7.0] |
| Other* | 92 (69.7) | 6.0 [4.0; 7.0] |

\*Other are described in Supplementary table 1

*S. aureus* as a single pathogen or in association with other pathogens was more involved in brain-injured patients VAP than in non-brain-injured patients (difference: 33.1 [19.3; 46]). Conversely, Gram-negative bacteria were less present in brain-injured patients (difference: – 15.9 [-25.1; –6.2]), especially group 3 *Enterobacteriaceae* (difference: –11.1 [-20; –1.8]). Details in **table 4**. No situation or risk factor was identified to distinguish VAP involving *S. aureus* as a single pathogen from VAP involving *S. aureus* as a copathogen. Details in **table 5**.

**Table 4:**
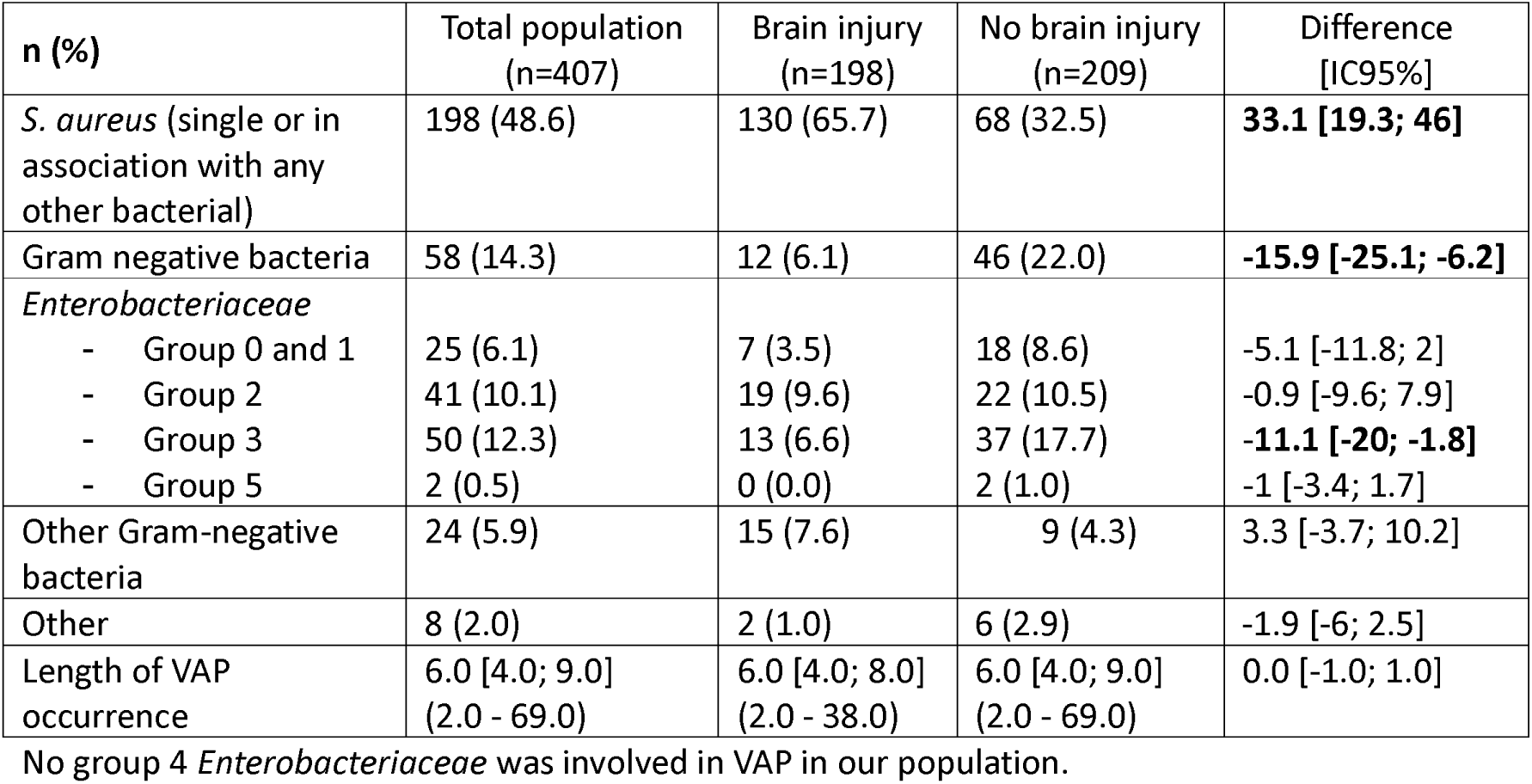
Comparison of microbiological specificities function of brain injury.

**Table 5:** Staphylococcus aureus VAP: factors associated with single vs multiple pathogen pneumonia.

| Variable<br>n (%) or mean± SD or P50<br>[P25; P75] | *S. aureus* VAP<br>(n=185) | Single pathogen (*S. aureus*) VAP<br>(n=53) | Multiple<br>pathogens VAP<br>(including *S. aureus*)<br>(n=132) | Difference<br>[IC95%] |
| --- | --- | --- | --- | --- |
| Sex |  |  |  |  |
| - Male | 125 (67.6) | 32 (60.4) | 93 (70.5) | 10.1 [-11.7; 32.1] |
| - Female | 60 (32.4) | 21 (39.6) | 39 (29.5) |  |
| Age (years) | 52 [36; 63] | 55 [45; 64] | 52 [35; 63] | 3.5 [-3.0; 8.0] |
| BMI (kg/m<sup>2</sup>) | 25.20 [22.5; 29.4] | 26.95 [22.3; 29.9] | 25.0 [22.5; 29.4] | 2.0 [-1.7; 3.2] |
| Hypertension | 35 (19.0) | 11 (20.8) | 24 (18.3) | 2.4 [-15.2; 22] |
| Atrial fibrillation | 4 (2.2) | 1 (1.9) | 3 (2.3) | -0.4 [-6.5; 9.6] |
| Chronic heart failure (no) | 180 (97.8) | 53 (100.0) | 127 (96.9) | 3.1 [-5.9; 7.6] |
| Arteritis | 0 (0.0) | 0 (0.0) | 0 (0.0) | 0 [-2.8; 6.7] |
| Smoking habits (yes) | 53 (32.5) | 16 (34.0) | 37 (31.9) | 2.1 [-20.3; 25.8] |
| COPD | 3 (1.6) | 1 (1.9) | 2 (1.5) | 0.4 [-5.4; 9.9] |
| Chronic respiratory failure | 0 (0.0) | 0 (0.0) | 0 (0.0) | 0 [-2.8; 6.7] |
| Long term oxygen therapy | 0 (0.0) | 0 (0.0) | 0 (0.0) | 0 [-2.8; 6.7] |
| Diabetes melitus |  |  |  |  |
| - No | 163 (88.6) | 45 (84.9) | 118 (90.1) | -5.2 [-22.2; 9.6] |
| - Uncomplicated | 14 (7.6) | 6 (11.3) | 8 (6.1) | 5.2 [-7.4; 20.4] |
| - Complicated | 7 (3.8) | 2 (3.8) | 5 (3.8) | 0 [-8.2; 11.7] |
| Chronic kidney disease |  |  |  |  |
| - None | 177 (96.2) | 52 (98.1) | 125 (95.4) | 2.7 [-8.4; 9.7] |
| - Without dialysis | 4 (2.2) | 0 (0.0) | 4 (3.1) | -3.1 [-7.6; 5.9] |
| - Requiring chronic dialysis | 3 (1.6) | 1 (1.9) | 2 (1.5) | 0.4 [-5.4; 9.9] |
| Alcohol abuse | 35 (21.5) | 10 (21.3) | 25 (21.6) | -0.3 [-19.4; 21.2] |
| Cirrhosis | 0 (0.0) | 0 (0.0) | 0 (0.0) | 0 [-2.8; 6.7] |
| Brain injury | 122 (65.9) | 33 (62.3) | 89 (67.4) | -5.2 [-27.4; 16.5] |
| - Trauma | 48 (39.3) | 13 (39.4) | 35 (39.3) | 0.1 [-27.3; 28.7] |
| - SAH | 37 (30.3) | 12 (36.4) | 25 (28.1) | 8.3 [-18.2; 35.8] |
| - Ischemic stroke | 10 (8.2) | 0 (0.0) | 10 (11.2) | -11.2 [-19.7; 5.1] |
| - Haemorrhagic stroke | 27 (22.1) | 8 (24.2) | 19 (21.3) | 2.9 [-20.2; 28.9] |
| - Other | 0 (0.0) | 0 (0.0) | 0 (0.0) | 0 [-4.1; 10.6] |
| Glasgow coma score | 8 [5; 15]<br>(3 - 15) | 8 [6; 15]<br>(3 - 15) | 8 [5; 14]<br>(3 - 15) | 0.0 [-1.0; 7.0] |
| No brain injury |  |  |  |  |
| - Sepsis / septic shock | 2 (3.2)<br>1 (1.6) | 0 (0.0)<br>1 (5.0) | 2 (4.7)<br>0 (0.0) | -4.7 [-15.8; 16.3]<br>5 [-8.1; 24.9] |
| - Acute exacerbation of chronic organ failure | 6 (9.5)<br>9 (14.3)<br>41 (65.1) | 1 (5.0)<br>1 (5.0)<br>16 (80.0) | 5 (11.6)<br>8 (18.6)<br>25 (58.1) | -6.6 [-25; 21]<br>-13.6 [-33.3; 16.5]<br>21.9 [-16.6; 52.1] |
| - Planned surgery |  |  |  |  |
| - Trauma |  |  |  |  |
| - Acute respiratory failure |  |  |  |  |
| SAPS II | 46 [36; 55] | 47 [39; 60] | 46 [36; 55] | 1.5 [-3.0; 8.5] |
| Facial trauma | 30 (16.2) | 10 (18.9) | 20 (15.2) | 3.7 [-13; 22.5] |
| Chest trauma | 28 (15.1) | 6 (11.3) | 22 (16.7) | -5.3 [-19.9; 12.3] |
| Immunocompromission |  |  |  |  |
| - None | 168 (91.3) | 47 (88.7) | 121 (92.4) | -3.7 [-19.3; 9.3] |
| - HIV infection | 0 (0.0) | 0 (0.0) | 0 (0.0) | 0 [-2.8; 6.7] |
| - Long-term corticosteroid | 4 (2.2) | 2 (3.8) | 2 (1.5) | 2.2 [-4.9; 12.8] |
| - Hematological malignancy | 4 (2.2) | 1 (1.9) | 3 (2.3) | -0.4 [-6.5; 9.6] |
| - Solid tumor | 9 (4.9) | 4 (7.5) | 5 (3.8) | 3.7 [-6.6; 17] |
| - Immunosuppressive treatment | 3 (1.6) | 0 (0.0) | 3 (2.3) | -2.3 [-6.5; 6.2] |
| Length of stay before VAP | 6 [4; 8] | 6 [4; 8] | 6 [4; 8] | 0.0 [-1.0; 1.5] |
| Delay of initial intubation (days) | 1 [1; 1] | 1 [1; 5] | 1 [1; 1] | 0.0 [0.0; 0.0] |
| Blood transfusion before VAP |  |  |  |  |
| - None | 65 (52.8) | 25 (78.1) | 40 (44.0) | 34.2 [5.3; 57.2] |
| - 3 or less | 39 (31.7) | 4 (12.5) | 35 (38.5) | -26 [-45.7; 0.5] |
| - > 3 units of red blood cells | 19 (15.4) | 3 (9.4) | 16 (17.6) | -8.2 [-25; 14.6] |
| Previous antibiotic therapy* |  |  |  |  |
| - None | 129 (70.1) | 32 (60.4) | 97 (74.0) | -13.7 [-35.3; 7.9] |
| - Beta-lactamines | 55 (29.9) | 21 (39.6) | 34 (26.0) | 13.7 [-7.9; 35.3] |
| - Glycopeptides | 0 (0.0) | 0 (0.0) | 0 (0.0) | 0 [-2.8; 6.7] |
| - Aminosides | 0 (0.0) | 0 (0.0) | 0 (0.0) | 0 [-2.8; 6.7] |
| - Other | 0 (0.0) | 0 (0.0) | 0 (0.0) | 0 [-2.8; 6.7] |
| Pathogen |  |  |  |  |
| - *S. aureus* | 185 (100.0) | 53 (100.0) | 132 (100.0) | - |
| - *E. coli* | 24 (13.0) | 0 (0.0) | 24 (18.2) | -18.2 [-25.8; -5.3] |
| - *E. cloacae* | 9 (4.9) | 0 (0.0) | 9 (6.8) | -6.8 [-12.5; 3.6] |
| - *C. koserii* | 12 (6.5) | 0 (0.0) | 12 (9.1) | -9.1 [-15.3; 1.9] |
| - *K. pneumoniae* | 21 (11.4) | 0 (0.0) | 21 (15.9) | **-15.9 [-23.3; -3.4]** |
| - *E.aerogenes* | 15 (8.1) | 0 (0.0) | 15 (11.4) | -11.4 [-18; 0.2] |
| - *P. aeruginosa* | 11 (5.9) | 0 (0.0) | 11 (8.3) | -8.3 [-14.4; 2.5] |
| - Other** | 92 (49.7) | 0 (0.0) | 92 (69.7) | -69.7 [-77.4; -54.4] |
| Septic shock | 71 (39.0) | 23 (43.4) | 48 (37.2) | 6.2 [-16.3; 28.9] |
| Leukocytes (G/l) | 13.0 [9.7; 17.6] | 12.3 [9.12; 15.9] | 13.2 [10.3; 18.0] | -0.8 [-3.3; 0.7] |

BMI : Body mass index; VAP: Ventilator-associated pneumonia
\* Previous antibiotic therapy during ICU stay
\*\* Detail of pathogens associated with *S. aureus* in **supplementary table 2**

### Outcomes

Median length of ICU stay was 25 [15; 39] days in the whole population. Specific length of stay was 27 [15; 40] in brain-injured population and 24 [14; 36] in patient without brain insult. Length of mechanical ventilation was 20 [13; 32] among the whole population, 21 [13; 32] and 20 [12; 33] in the two subpopulations. Details in **table 6**. A total of 172 patients (42.3%) died during their ICU stay and 90-days mortality was 194 (47.7%). Details in **table 6**.

**Table 6:** outcomes.

|  | Total population<br>(n=407) | Brain-injured<br>patients<br>(n =198) | Patients without<br>brain injury<br>(n= 209) |
| --- | --- | --- | --- |
| Length of ICU stay; median [IQ 25; 75]) | 25 [15; 39] | 27 [15; 40] | 24 [14; 36] |
| Length of invasive mechanical<br>ventilation; median [IQ 25; 75]) | 20 [13; 32] | 21 [13; 32] | 20 [12; 33] |
| Mortality |  |  |  |
| - D28 mortality; n (%) | 141 (34.6) | 57 (28.8) | 84 (40.2) |
| - ICU mortality; n (%) | 172 (42.3) | 70 (35.5) | 102 (48.8) |
| - D90 mortality; n (%) | 194 (47.7) | 77 (38.9) | 117 (55.9) |
| - Length between admission and<br>death ; median [IQ 25; 75]) | 17 [11; 29] | 15 [10; 29] | 18 [13; 31] |

The comparison of mortality in function of the involvement of *S. aureus* during VAP did find any difference, neither in brain-injured patients (p= 0.661; cf. **figure 2**) nor in non-brain injured patients (p= 0.572; cf. **figure 3**).

### Risk factor of involvement of *S. aureus* in VAP

In the whole population, univariate analysis identified age (0.96 [0.94; 0.97]; p < 0.0001), chronic heart failure (0.27 [0.08; 0.74]; p = 0.0092), arteritis (p< 0.0001), COPD (0.25 [0.06; 0.77]; p = 0.0137), chronic renal failure (0.22 [0.06; 0.6]; p = 0.0077), higher Glasgow coma score (0.88 [0.85; 0.92]; p < 0.0001), SAPS II (0.98 [0.97; 0.99]; p = 0.0024), previous antibiotic therapy by beta-lactamins (0.56 [0.37; 0.84]; p = 0.0048) as protective factors, and brain injury (3.72 [2.48; 5.64]; p < 0.0001), especially, facial trauma (2.07 [1.13; 3.87]; p = 0.0182), intracranial pressure monitoring (p < 0.0001), initial hemorrhagic shock (2.36 [1.23; 4.69]; p=0.0096) as risk factors for involvement of *S. aureus* in VAP. Detail in **table 7**.

**Table 7:**
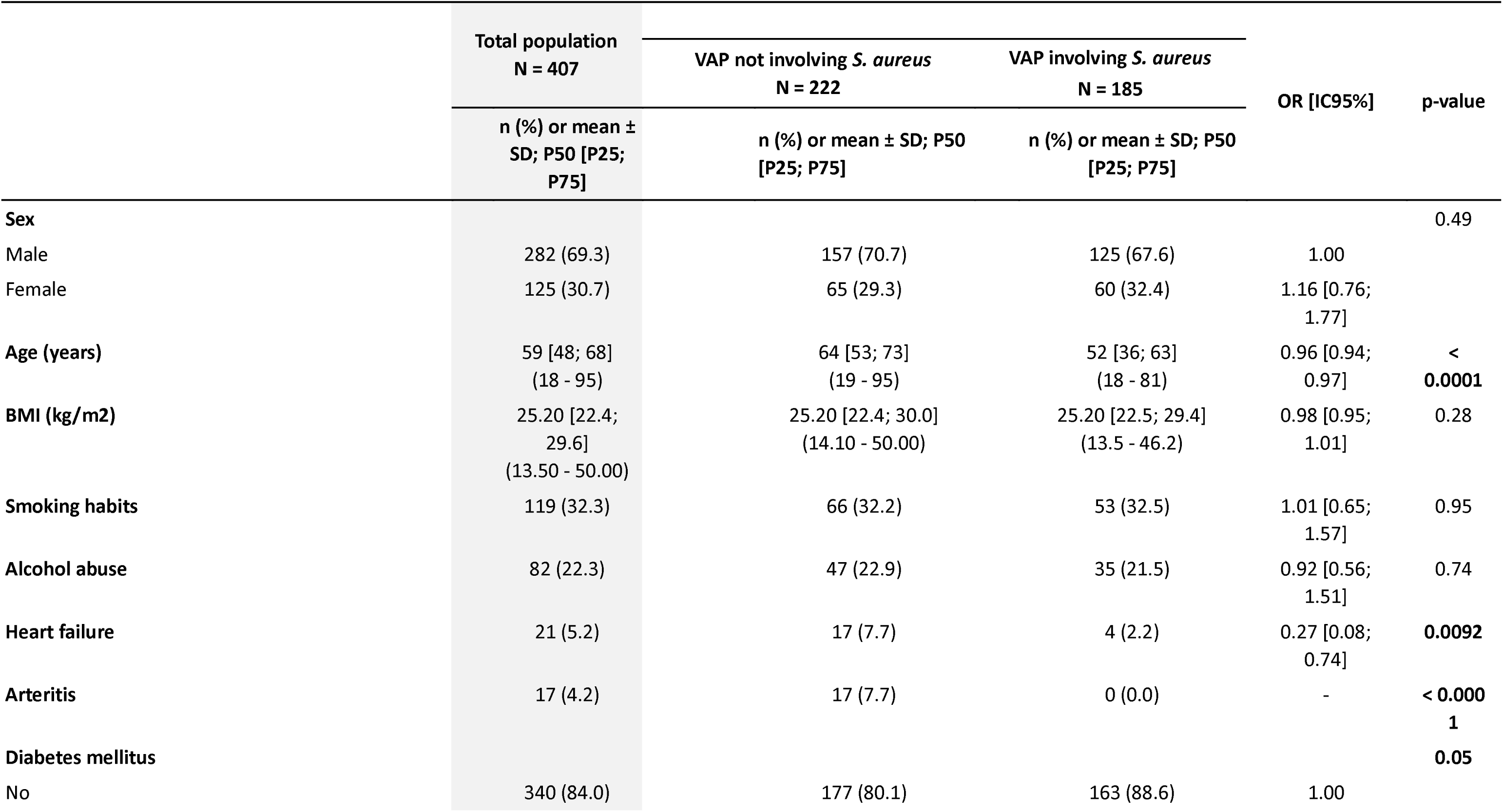
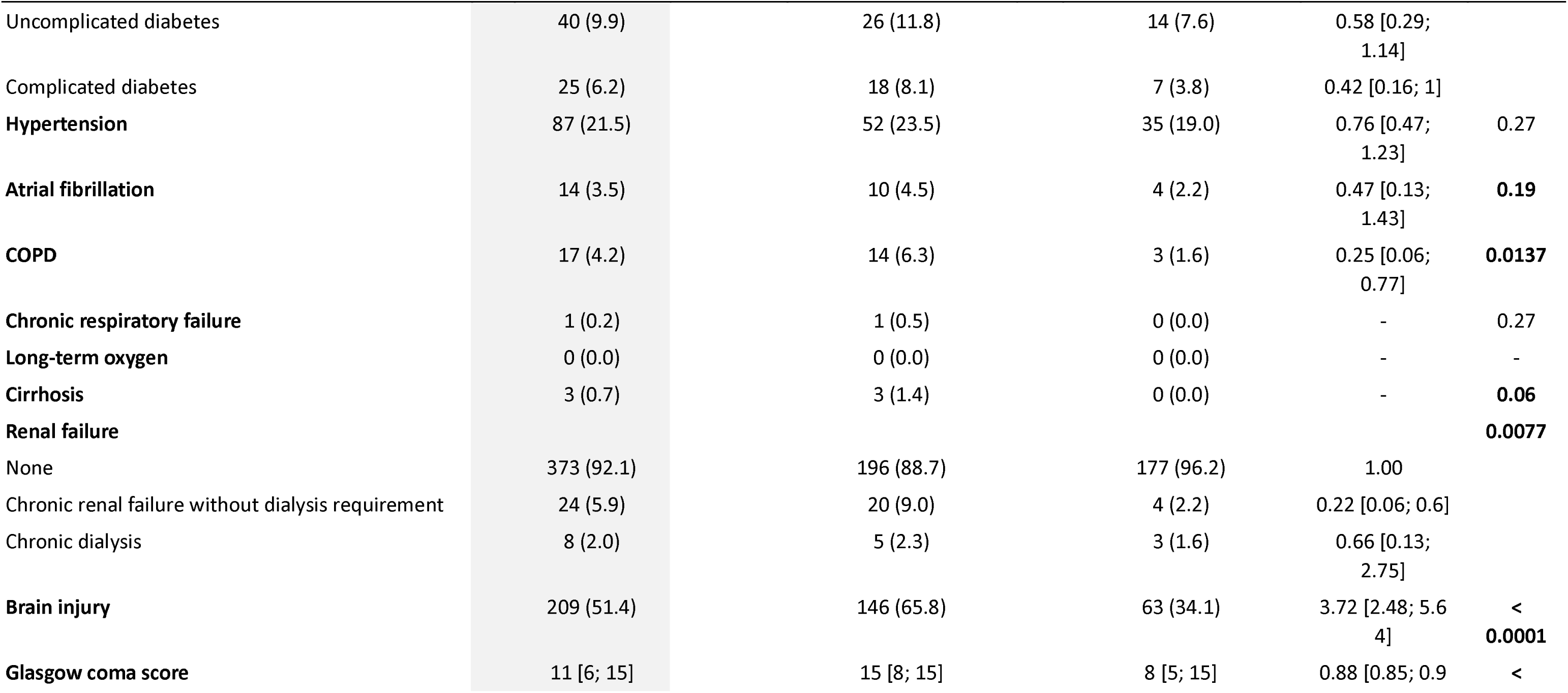
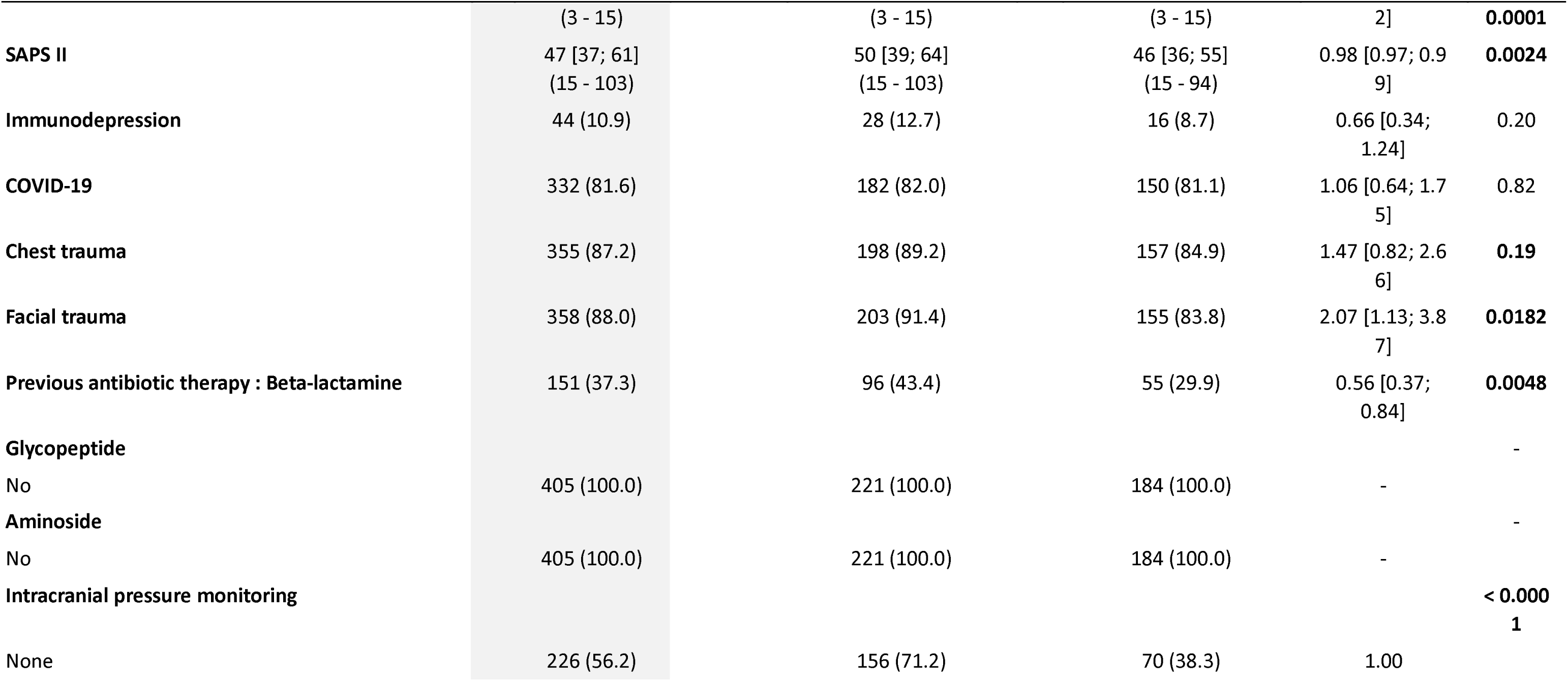
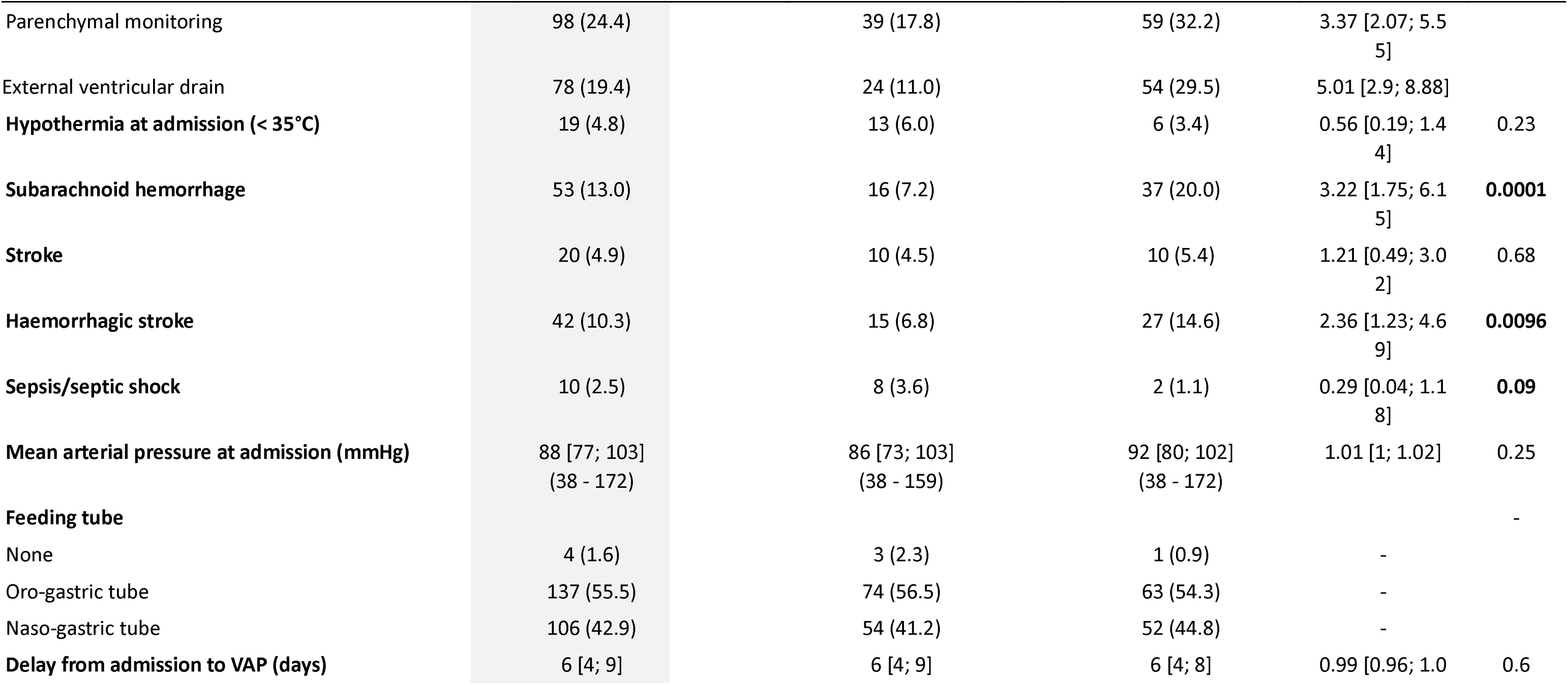
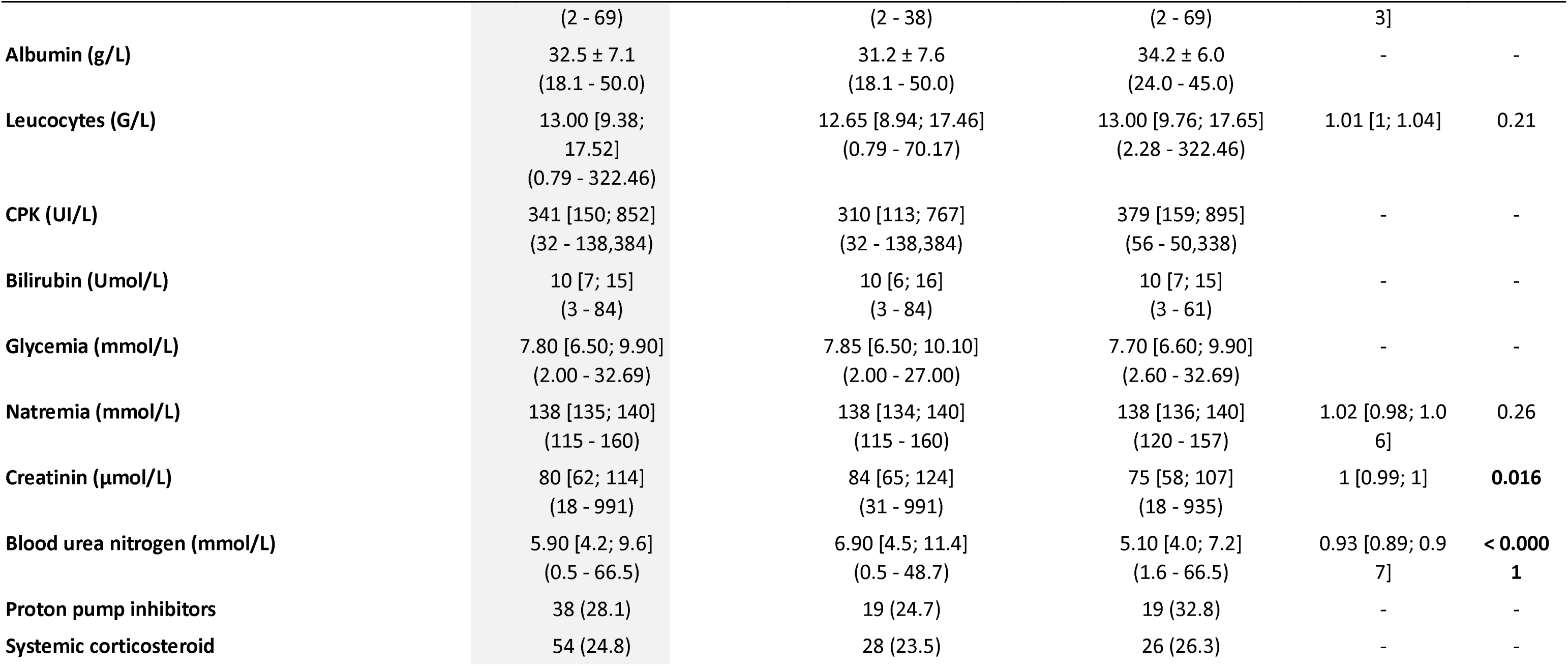
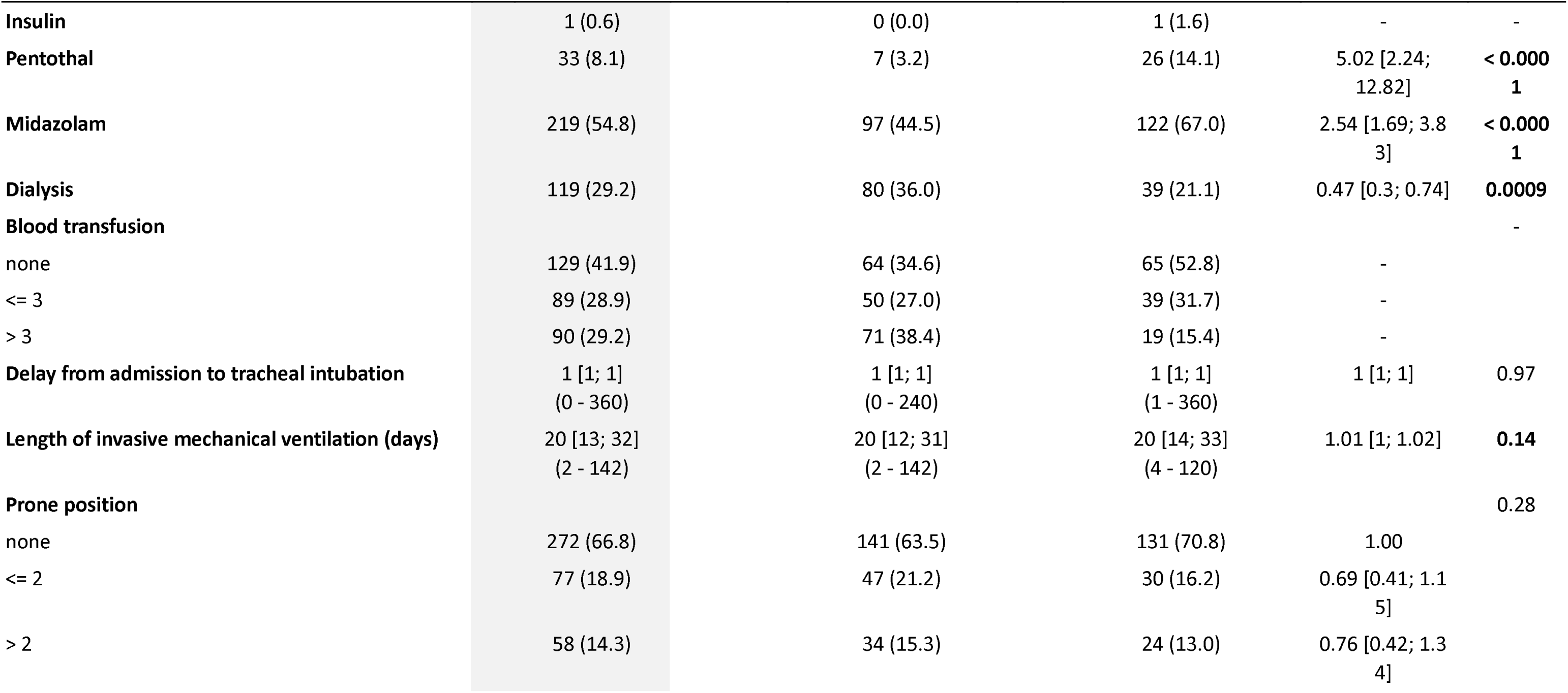
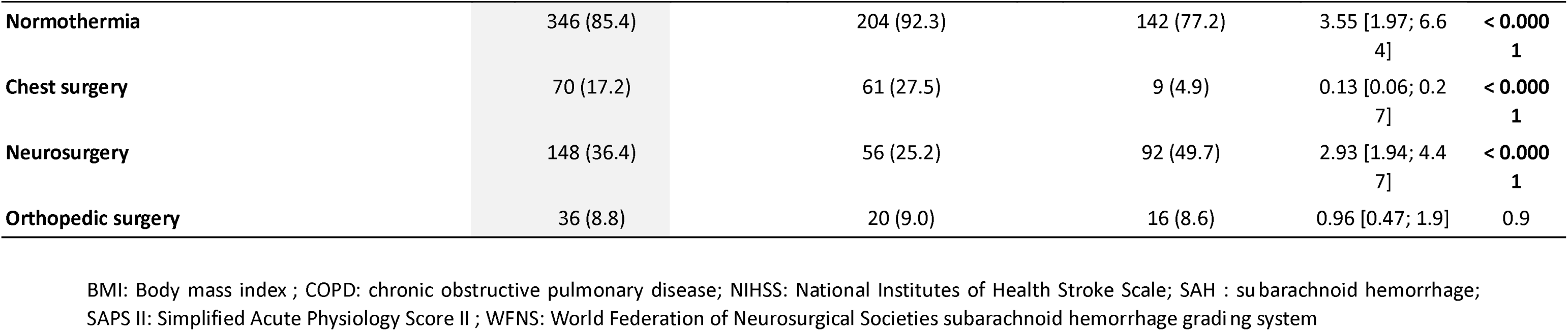
risk factors associated with S. aureus VAP in the whole population.

The multivariate analysis identified thoracic surgery (absence: 0.26 [0.11; 0.59]; p=0.0019), and lower Glasgow coma score (0.94 [0.88; 1]; p=0.0458) p=0.0458) as risk factors. Conversely, previous antibioprophylaxis (in the initial surgical context) (0.36 [0.19; 0.65]; p=0.0009), age (0.97 [0.96; 0.98]; p<0.0001), and SAPS II (0.98 [0.97; 1]; p=0.0151) were protective factors. Details in **table 8** and **supplementary table 4** and **supplementary figure 1**.

**Table 8:** risk factors of involvement of Staphylococcus aureus in VAP in the whole population: multivariate analysis.

| Variable (reference) | OR [IC95%] | p-value |
| --- | --- | --- |
| Age (years) | 0.97 [0.96; 0.98] | < 0.0001 |
| Per-surgery antibioprophylaxis | 0.36 [0.19; 0.65] | **0.0009** |
| Thoracic surgery | 0.26 [0.11; 0.59] | **0.0019** |
| SAPS II | 0.98 [0.97; 1] | **0.0151** |
| Glasgow coma score | 0.94 [0.88; 1] | **0.0458** |
| Previous treatment with beta-lactamins | 0.65 [0.37; 1.14] | 0.13 |
| Subarachnoid hemorrhage | 1.96 [0.89; 4.59] | 0.11 |
| Cirrhosis (no) | - | > 0.99 |
| Length of mechanical ventilation (days) | 1.01 [1; 1.02] | 0.17 |
| Arteritis (no) | - | 0.99 |
| Chronic respiratory failure (no) | - | > 0.99 |

In brain-injured patients, multivariate analysis identified age (0.97 [0.95; 0.98]), antibioprophylaxis (0.31 [0.18; 0.53]; p= < 0.0001), subarachnoid hemorrhage (2.18 [1.01; 4.68]; p=0.0461), lower Glasgow coma score (0.92 [0.87; 0.97]; p= 0.004) and SAPS II (0.98 [0.97; 1]; p=0.0174) as critical factors associated with the risk of *S. aureus* in VAP. Details in **table 9 and supplementary table 5 and 6.** In the non-brain-injured population, multivariate analysis identified antibioprophylaxis (0.38 [0.22; 0.65]; p=0.0006), age (0.96 [0.95; 0.98]; p<0.0001), SAPS II (0.98 [0.97; 1]; p=0.0265), thoracic surgery (0.21 [0.09; 0.48]; p=0.0002) and peripheral neurosurgery (1.95 [1.2; 3.19]; p=0.0076) as critical factors. Details in **table 10** and **supplementary table 7 and 8**.

**Table 9:**
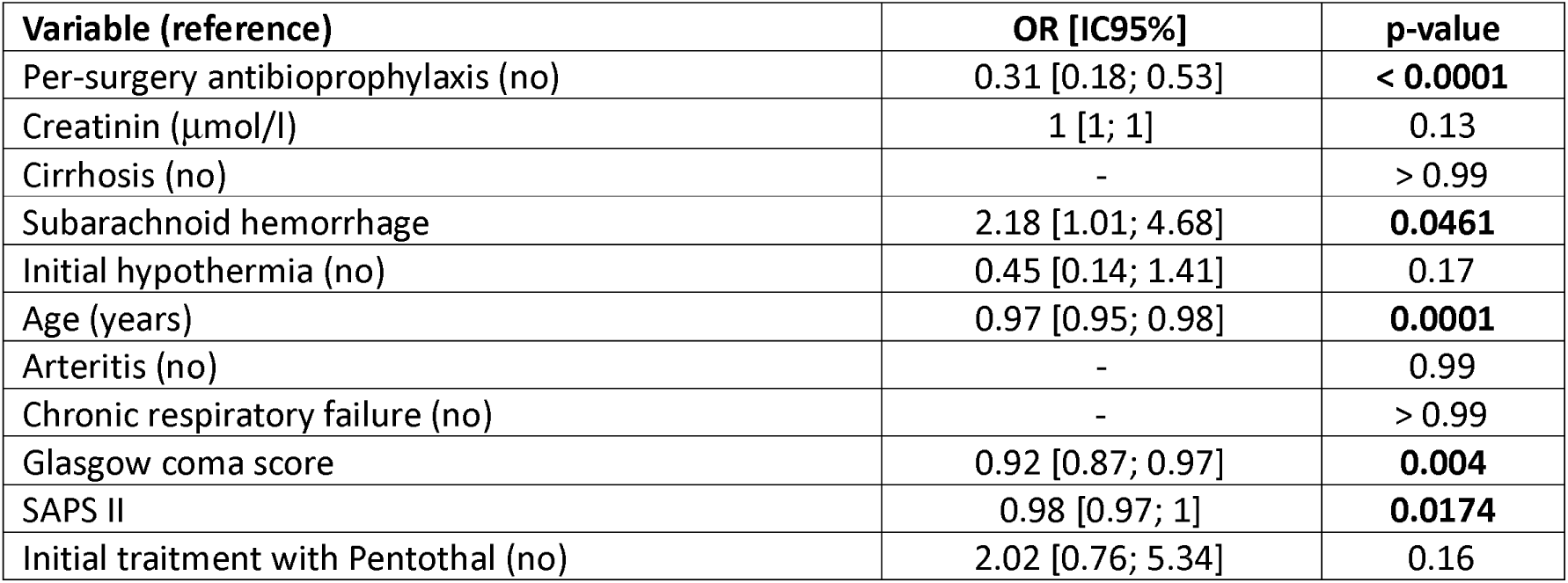
Subgroup analysis: risk factors of involvement of Staphylococcus aureus in VAP in brain-injured population.

| Variable (reference) | OR [IC95%] | p-value |
| --- | --- | --- |
| Per-surgery antibioprophylaxis (no) | 0.31 [0.18; 0.53] | < 0.0001 |
| Creatinin (μmol/l) | 1 [1; 1] | 0.13 |
| Cirrhosis (no) | - | > 0.99 |
| Subarachnoid hemorrhage | 2.18 [1.01; 4.68] | **0.0461** |
| Initial hypothermia (no) | 0.45 [0.14; 1.41] | 0.17 |
| Age (years) | 0.97 [0.95; 0.98] | **0.0001** |
| Arteritis (no) | - | 0.99 |
| Chronic respiratory failure (no) | - | > 0.99 |
| Glasgow coma score | 0.92 [0.87; 0.97] | **0.004** |
| SAPS II | 0.98 [0.97; 1] | **0.0174** |
| Initial treatment with Pentothal (no) | 2.02 [0.76; 5.34] | 0.16 |

**Table 10:** Subgroup analysis: risk factors of involvement of Staphylococcus aureus in VAP in non-brain-injured population.

| Variable (reference) | OR [IC95%] | p-value |
| --- | --- | --- |
| Per-surgery antibioprophylaxis (no) | 0.38 [0.22; 0.65] | **0.0006** |
| Age (years) | 0.96 [0.95; 0.98] | < 0.0001 |
| Arteritis (no) | - | 0.99 |
| SAPS II | 0.98 [0.97; 1] | **0.0265** |
| Thoracic surgery | 0.21 [0.09; 0.48] | **0.0002** |
| Neurosurgery | 1.95 [1.2; 3.19] | **0.0076** |
| Length of invasive mechanical ventilation | 1.01 [1; 1.02] | 0.17 |

## Discussion

In our cohort of 407 patients, all patients developed a VAP. No difference was noticed in time to VAP occurrence between groups. *S. aureus* was the predominant pathogen in both groups of patients, although its prevalence was higher among brain injured patients. Conversely, Gram-negative bacteria, especially group 3 *Enterobacteriaceae* were more frequent in non-brain injured patients. The association of *S. aureus* with other pathogens was similar in both groups of patients and repartition of other pathogens was not different. Of note the most represented pathogen in association with S. aureus was *H. influenzae*. Factors identified as being associated with the risk of developing a VAP involving *S. aureus* in our brain-injured population was age, Glasgow coma score the initial subarachnoid hemorrhage and the severity, illustrated by SAPS II. In the non-brain-injured patients, age and SAPS II are also independent risk factors of *S. aureus*-associated VAP and are associated with peripheral neurosurgery and thoracic surgery as protective factors. In both groups, antibioprophylaxis is associated with a reduction risk of *S. aureus* involvement.

To the best of our knowledge, this is the first multicenter study to analyze risk factors for *S. aureus*-associated VAP by comparing two cohorts of patients, one with brain injury and one without. The importance of *S. aureus* in VAP among brain-injured patients is well established^59,60^. Its involvement in this context may be more than twice its incidence during VAP in patients without brain injury^9,10^, which is coherent with our observations (61.6% *vs* 30.1%).

The risk factors associated with *S. aureus* involvement in VAP are consistent with what has already been reported in the literature^61–65^.

The increased risk of infection in brain-injured patients is well known^66^. Numerous studies have focused on the risk of developing VAP following brain injury, identifying a variety of factors such as male sex^67,68^, smoking^67,69^, immunosuppression^70^, initial aspiration^71,72^, arterial hypertension^71^, intracranial hypertension^71^, barbiturate use^70,71^, mannitol^68^, hypothermia^67,72^, the presence of an intraparenchymal brain probe^67^, bronchial inflammation^67^, and the severity of both respiratory (ARDS)^70^ and systemic conditions (as illustrated by the SAPS II score)^70^. Notably, subarachnoid hemorrhage, which was identified as a risk factor for *S. aureus* involvement in our study, had previously been observed as a risk factor for VAP in a population where *S. aureus* was highly prevalent (34.9%)^68^.

The protective effect of surgery could be linked to systematic nasal decontamination during surgical preparation, or more generally to antibiotic prophylaxis, which is associated with a reduced risk of VAP in the context of brain injury^73^. Moreover, the latter is strongly associated with the absence of *S. aureus* involvement. In a study by Karim Asehnoune’s team, neurosurgery was also identified as a protective factor against the occurrence of pneumonia^70^. However, although *S. aureus* was widely represented during VAP in this population, the risk factors did not focus specifically on this pathogen. Coma depth or the SAPS II score, illustrating the severity of neurological or systemic impairment, likely represent markers of systemic alteration and impaired immune defenses. Similarly, age may reflect immunosenescence (although this factor remains debated^69,74^) or nutritional status, though we are unable to confirm this due to our study design

Although the involvement of *S. aureus* in VAP, particularly in cases of brain injury, has long been recognized^75–80^, the origin of this specificity remains poorly understood. The central element in the infectious risk among brain-injured patients is systemic immune impairment. Acute brain injury disrupts homeostasis by inducing an increase in intracranial pressure, cerebral ischemia, blood-brain barrier disruption, or cellular damage leading, in particular, to the release of damage-associated molecular patterns^81,82^. Concomitantly, a compensatory anti-inflammatory response leads to a state of immunosuppression^83^, described as CNS injury-induced immunosuppression (CIDS), which can promote the occurrence of infections^84^ without a clear pathogen distinction. The mechanisms responsible for this peripheral immune impairment appear to be centered on the autonomous nervous system and the hypothalamic-pituitary endocrine system^85^. Activation of these structures could lead to a reduction in the inflammatory response in favor of the production of mediators such as interleukin-10 or TGF-β, and result in the restriction of T-cell migration and proliferation^86^. The neuro-immunological axis could also promote lymphocyte apoptosis and impair macrophage activation^87,88^. In addition to this quantitative reduction, these phenomena are accompanied by a loss of cytotoxic activity and a shift from a Th1 polarity toward a more tolerant Th2 phenotype^84,89,90^. Furthermore, brain injury is associated with peripheral immune impairment, particularly pulmonary dysfunction in experimental models^91^, which tends to be confirmed in humans by the high incidence of respiratory infections^92^. Finally, the pulmonary capillary leak responsible for the exudative alveolar edema observed in brain injuries could also predispose patients to alveolar infectious risk. Unfortunately, none of these mechanisms are specifically associated with *S. aureus* or even Gram-positive cocci.

In addition to immunological changes that facilitate pathogen proliferation by weakening patient defenses, VAP are promoted by microaspiration through the endotracheal tube^93,94^, transcolonization^95,96^, and alterations of the respiratory microbiota. Changes occurring within the ENT sphere^97,98^ promote the subsequent migration of these pathogens into the subglottic space^99,100^. Nasal colonization by *S. aureus*, which is more frequent in younger individuals^101^, could also promote its proliferation within the oral cavity and oropharynx before allowing its passage into the subglottic space. The presence of identical strains in both the nasal cavities and respiratory samples supports this hypothesis^102^.

Of particular interest in this context is the significance of the interactions between *S. aureus* and *Candida albicans*. Indeed, *C. albicans* is widely present in the oral cavity. Furthermore, it is frequently found in the lower respiratory tract of patients during invasive mechanical ventilation^103,104^. Multiple studies have demonstrated a link between the presence of *Candida* in the respiratory tract and the risk of bacterial pneumonia in mechanically ventilated patients^105–107^. On the other hand, the presence of *C. albicans* in the airways is associated with the presence of *S. aureus*^108–110^. Although clinical evidence remains uncertain, available models confirm the existence of an interaction between *Candida* and several bacteria, particularly within the respiratory tract^111^. The elementary *Galleria mellonella* model demonstrates increased mortality when *C. albicans* is combined with *S. aureus*^112^. Furthermore, the administration of *C. albicans* in rats promotes the subsequent occurrence of *S. aureus* pneumonia^107^. This specific feature is underpinned by several interactive mechanisms between the two microbial agents. The adhesion of *S. aureus* to *C. albicans* hyphae appears to promote the production of a shared biofilm, driven notably by the production of prostaglandin E2 by *C. albicans*^113^. Additionally, *C. albicans* biofilm co-culture favors the secondary bacterial proliferation of *S. aureus*^112,114^. Contact with *S. aureus* is facilitated by Agglutinin-like sequence 3 (Als3), which binds to bacterial integrins, particularly fibronectin-binding protein B)^115^. Als3 also appears to play a role in *S. aureus* proliferation within the polymicrobial biofilm, at least during the early phase^116^. Additionally, *C. albicans* may enhance *S. aureus* virulence by upregulating the activation of the Agr system, which in turn leads to increased alpha-toxin production^117^. Finally, the interaction between *Candida* and *S. aureus* drives numerous gene alterations, upregulating the expression of metabolic genes, virulence factors, and defense mechanisms against host responses, particularly oxidative stress^118^.

Our study has several limits. First, due to its retrospective design, our study is subject to data collection bias; however, the combined search across electronic health files, microbiology and laboratory and image software yielded comprehensive data for all included patients. Similarly, potential recall bias that could have reduced the number of admitted patients was bypassed by utilizing the coding system (based on the ICD-10 classification of diseases) required for hospital funding. The lack of a standardized definition for VAP was mitigated by the consistent use of the same criteria across all participating centers reducing the risk of heterogeneity between centers. Second, the study design did not allow for an assessment of VAP incidence during the ICU stay, particularly within the brain-injured patient subgroup, nor for the evaluation of risk factors for infection onset; however, these aspects have already been extensively investigated^24,119^. Third, the absence of patients who did not develop VAP precluded an assessment of the impact of *S. aureus*-associated pneumonia on ICU mortality. However, VAP-associated mortality remains challenging to evaluate and has already been the subject of numerous publications. Furthermore, in our study, comparing patients with VAP involving *S. aureus* to those infected with other pathogens revealed no significant differences between the groups. Similarly, the lack of data on the initial efficacy of antibiotic therapy during VAP leaves the effect on mortality uncertain; however, the absence of methicillin-resistant *S. aureus* carriage and its absence during pneumonia suggest that the treatment was effective against this pathogen. Moreover, methicillin-resistant-*S. aureus* are already known to play a minor role during early VAP^9,120^. Finally, due to the retrospective nature of the study, we were unable to obtain more precise information regarding the immunological changes promoting the occurrence of pneumonia specifically *S. aureus* pneumonia, or the alterations in the microbiota and mycobiota that could favor this pathogen, particularly in brain-injured patients. Our study has also several strengths. First, its multicenter design, which eliminates the influence of center-specific microbial ecology and local clinical practices, ensures robust external validity. Second, the large sample size provided sufficient statistical power to identify several risk factors for *S. aureus* involvement during VAP, allowing us to differentiate these factors based on the presence or absence of brain injury. Similarly, we were able to categorize brain injuries according to their specific type. Third, by comparing patients admitted for brain injury against those with other pathologies, we were able to compare the microbiological epidemiology and confirm the overrepresentation of S. aureus among VAP pathogens in brain-injured patients. At last, the absence of methicillin-resistant *S. aureus* in our cohort allowed us to investigate the presence and impact of S. aureus while adjusting for confounding factors related to the efficacy of empirical antibiotic coverage or lower respiratory tract colonization driven by nasal carriage.

## Conclusion

In our multicenter study, the involvement of S. aureus is increased in the brain-injured group of patients. Factors identified as being associated with the risk of developing a VAP involving *S. aureus* in our brain-injured population was age, Glasgow coma score the initial subarachnoid hemorrhage and the severity, illustrated by SAPS II. In both groups, antibioprophylaxis is associated with a reduction risk of *S. aureus* involvement.

## Supporting information

Supplementary data set

## Acknowledgement

none

## Authors contribution

Conceptualization: LF, FP; Investigation: LF, CS,NM,CQ, FP; Data curation: LF, MY; Methodology: MY; Formal analysis: MY; Validation: MY, LF, FP; Supervision: FP; Writing original draft: LF, FP; Writing review and editing: CS, NM, CQ, FP

## Confict of interest

none

## Data availability

Datasets used/analyzed in this study are accessible upon reasonable request to the corresponding author.

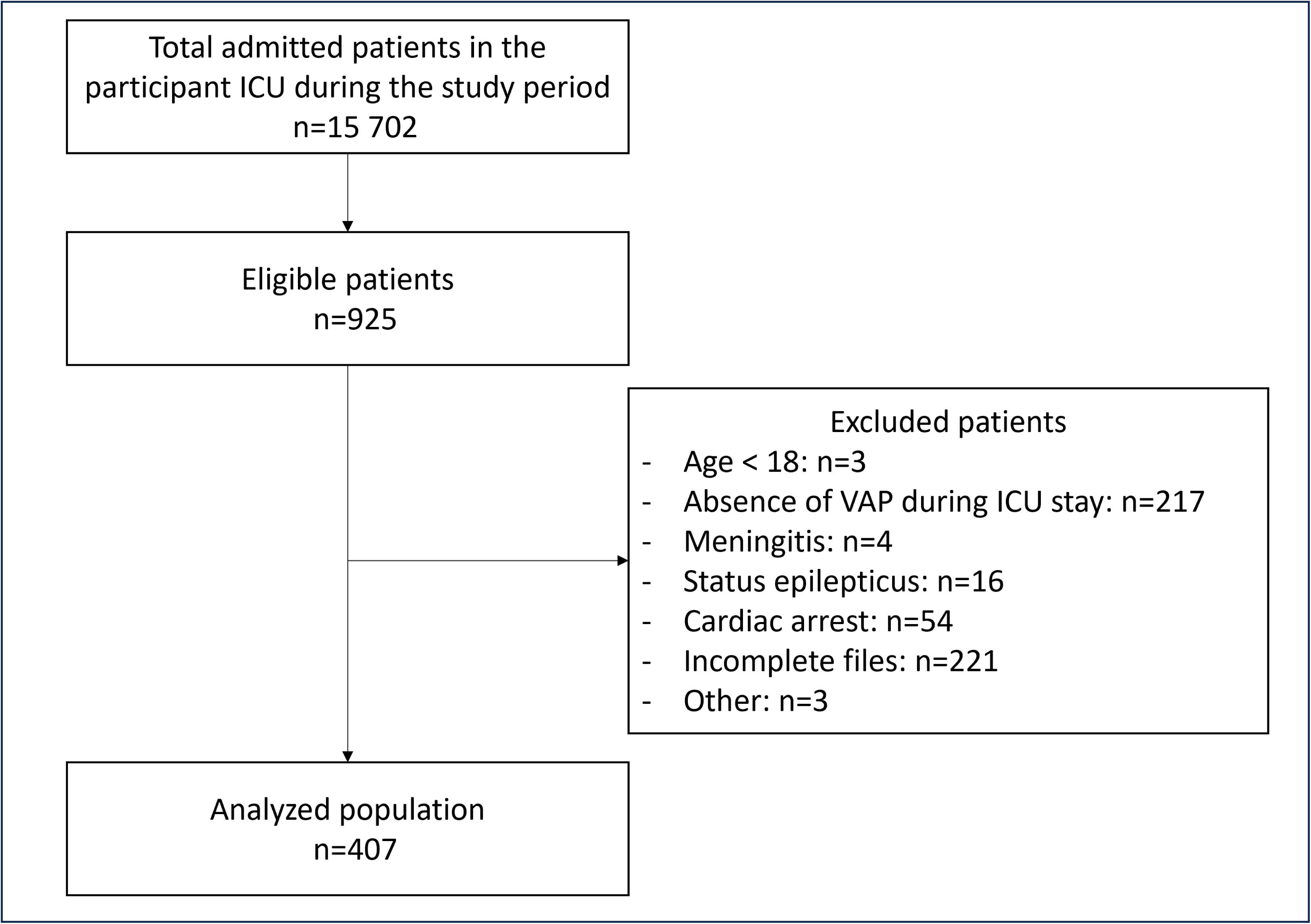

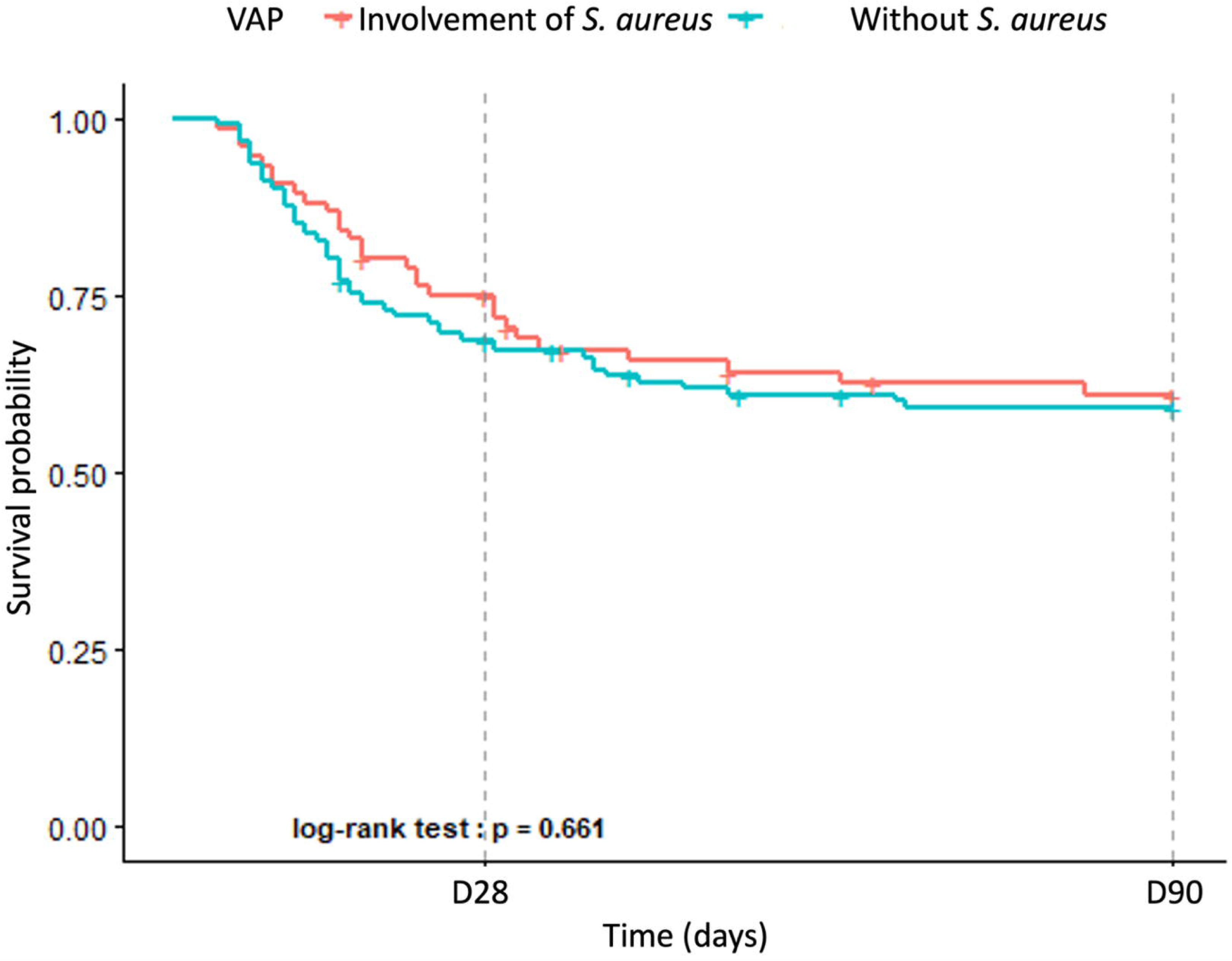

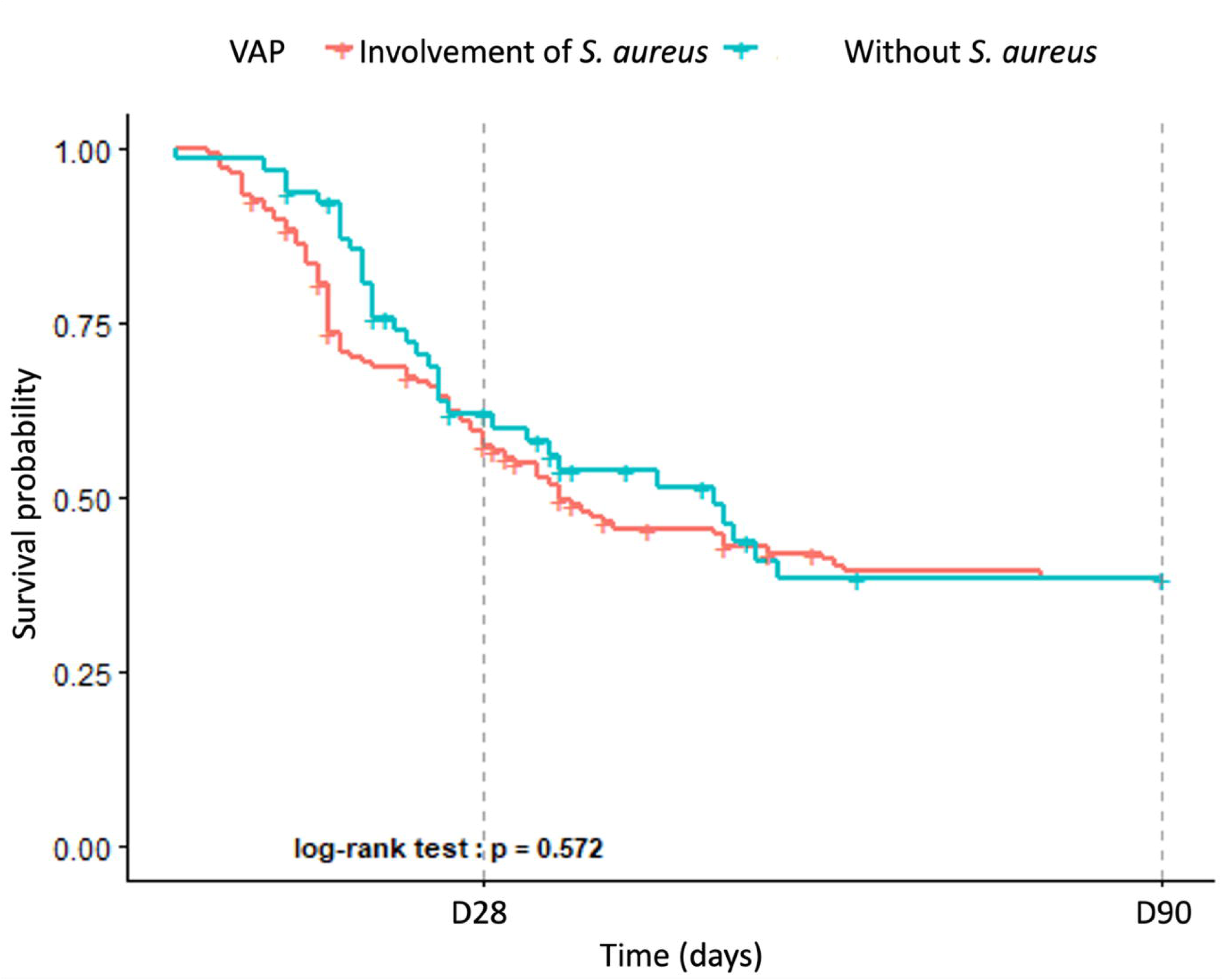

