## Supplementary data set for "Microbiological characteristics of ventilator-associated pneumonia in patients with neurological injuries: a multicentric cohort"

### Supplementary material

#### Statistical analysis

Quantitative variables were presented as means (+/- standard deviation) or medians [interquartile range] and minimum-maximum, depending on their distribution, evaluated using visual inspection and normality tests. Qualitative variables were described in terms of numbers and percentages. When appropriate, differences in means, medians, or proportions between groups were reported with 95% confidence intervals (95% CI), computed using normal approximation or bootstrap methods for means and medians, and the Clopper-Pearson method for proportions.

Missing data: Variables with more than 20% missing data or with a single observed category were excluded from the candidate list and were not imputed. Admission-diagnosis variables that were structurally inapplicable to a given population (i.e., neurological diagnoses in non-brain-injured patients, and sepsis/septic shock in brain-injured patients) were coded as « no » rather than imputed. Missing data were assumed to be missing at random (MAR) and were handled using multiple imputation by chained equations (MICE), generating 40 imputed datasets (20 iterations). Predictive mean matching, logistic regression, and polytomous logistic regression were used for continuous, binary, and multi-category variables, respectively. Convergence was assessed graphically.

Univariate analysis: A list of candidate risk factors was defined a priori. Odds ratios (ORs) with their 95% CI were computed on the original data, and p-values were calculated using the likelihood ratio test. Variables with a p-value $< 0.20$ were selected. No variables were forced into the model.

Multivariate analysis: The final multivariate model was obtained using a backward stepwise selection procedure, applied independently to each of the 40 imputed datasets to account for imputation-related uncertainty during the variable selection process. For each candidate variable, the selection frequency across the 40 replications was calculated. A majority rule was then applied: only variables selected in at least 50% of the imputed datasets were retained in the final model. The resulting model was subsequently refit to each imputed dataset, and the coefficient estimates were pooled using Rubin's rules to derive adjusted odds ratios and their 95% CI.

Internal validation: Optimism correction was performed using a bootstrap procedure adapted to multiply imputed data (200 replications), with the complete modelling process (univariate screening, backward stepwise selection, model fitting, pooling using Rubin's rules) repeated at each replication. The same resampling indices were applied identically across the 40 imputed datasets. Model performance was assessed both on the bootstrap and original imputed datasets, and optimism was estimated as the average difference between these two performances across replications. Discrimination was assessed using the c-statistic, and calibration using the calibration slope and the observed-to-expected (O/E) ratio; Cox and Snell's $R^2$ was used as a measure of explained variation.

Sensitivity analysis: A sensitivity analysis was performed using complete-case analysis, to assess the robustness of the ORs from the multiple imputation-based model.

Subgroup analysis: A pre-specified subgroup analysis was performed according to neurological injury status (brain-injured vs. non-brain-injured patients). For each subgroup, the same modelling strategy as the main analysis was reproduced independently. In the brain-injured subgroup, additional candidate variables specific to this population were included.

Exploratory analyses: Survival time was calculated from the date of ICU admission to death or, if the patient survived, up to day 90 post-admission. Patients with an unknown vital status at day 90 were censored at the date of last follow-up, defined as the most recent date between ICU discharge and day 28 post-ICU admission. Survival curves were compared using the log-rank test.

The significance threshold has been set at $p < 0.05$. Statistical analysis were performed using the R® software (R v4.4.2, URL: https://www.R-project.org/).

**Supplementary table 1**: initial characteristics of brain-injured patients

| Variable | Brain-injured patients N = 198 | Type of injury | | | | |
|---|---|---|---|---|---|---|
| | | Trauma N = 79 | Subarachnoid hemmorhage N = 53 | Ischemic stroke N = 20 | hematoma N = 42 | Other N = 4 |
| | n (%) or mean ± sd ; P50 [P25 ; P75] | n (%) or mean ± sd ; P50 [P25 ; P75] | n (%) or mean ± sd ; P50 [P25 ; P75] | n (%) or m ean± sd ; P50 [P25 ; P75] | n (%) ou mean ± sd ; P50 [P25 ; P75] ; | n (%) ou mean ± sd ; P50 [P25 ; P75] |
| **Sex** | | | | | | |
| Male | 134 (67.7) | 67 (84.8) | 27 (50.9) | 13 (65.0) | 25 (59.5) | 2 (50.0) |
| Female | 64 (32.3) | 12 (15.2) | 26 (49.1) | 7 (35.0) | 17 (40.5) | 2 (50.0) |
| **Age (years)** | 52 [36 ; 63] | 44 [27 ; 59] | 56 [49 ; 67] | 47 [35 ; 59] | 54 [41 ; 63] | 58 [53 ; 69] |
| **BMI (kg/m2)** | 24.00 [21.17 ; 27.80] (13.50 - 45.80) | 22.90 [20.30 ; 25.50] (13.50 - 39.50) | 24.69 [21.30 ; 29.38] (16.98 - 45.80) | 26.60 [22.90 ; 28.10] (18.52 - 37.90) | 24.84 [21.46 ; 28.34] (16.00 - 42.24) | 23.23 [21.10 ; 27.50] (21.10 - 27.50) |
| **No cardiac or vascular disease** | 144 (73.5) | 64 (83.1) | 35 (66.0) | 16 (80.0) | 27 (64.3) | 2 (50.0) |
| **Chronic heart failure** | | | | | | |
| No | 192 (98.0) | 75 (97.4) | 53 (100.0) | 19 (95.0) | 41 (97.6) | 4 (100.0) |
| yes | 4 (2.0) | 2 (2.6) | 0 (0.0) | 1 (5.0) | 1 (2.4) | 0 (0.0) |
| **Arteritis** | 2 (1.0) | 1 (1.3) | 1 (1.9) | 0 (0.0) | 0 (0.0) | 0 (0.0) |
| **Diabetes mellitus** | | | | | | |
| No | 175 (89.3) | 70 (90.9) | 45 (84.9) | 19 (95.0) | 38 (90.5) | 3 (75.0) |
| uncomplicated | 13 (6.6) | 4 (5.2) | 6 (11.3) | 1 (5.0) | 1 (2.4) | 1 (25.0) |
| Complicated | 8 (4.1) | 3 (3.9) | 2 (3.8) | 0 (0.0) | 3 (7.1) | 0 (0.0) |
| **Hypertension** | 29 (14.8) | 4 (5.2) | 10 (18.9) | 2 (10.0) | 12 (28.6) | 1 (25.0) |
| **Atrial fibrillation** | 3 (1.5) | 1 (1.3) | 1 (1.9) | 0 (0.0) | 1 (2.4) | 0 (0.0) |
| **COPD** | 5 (2.6) | 3 (3.9) | 1 (1.9) | 0 (0.0) | 1 (2.4) | 0 (0.0) |
| **Chronic respiratory failure** | 1 (0.5) | 0 (0.0) | 0 (0.0) | 0 (0.0) | 1 (2.4) | 0 (0.0) |
| **Long-term oxygen** | 0 (0.0) | 0 (0.0) | 0 (0.0) | 0 (0.0) | 0 (0.0) | 0 (0.0) |
| **Cirrhosis** | 1 (0.5) | 1 (1.3) | 0 (0.0) | 0 (0.0) | 0 (0.0) | 0 (0.0) |
| Chronic renal failure | 5 (2.6) | 1 (1.3) | 0 (0.0) | 4 (20.0) | 0 (0.0) | 0 (0.0) |

| | Brain-injured patients N = 198 | Type of injury | | | | |
|---|---|---|---|---|---|---|
| | | Trauma N = 79 | Subarachnoid hemmorhage N = 53 | Ischemic stroke N = 20 | hematoma N = 42 | Other N = 4 |
| **Variable** | **n (%) or mean ± sd ; P50 [P25 ; P75]** | **n (%) or mean ± sd ; P50 [P25 ; P75]** | **n (%) or mean ± sd ; P50 [P25 ; P75]** | **n (%) or m ean± sd ; P50 [P25 ; P75]** | **n (%) ou mean ± sd ; P50 [P25 ; P75] ;** | **n (%) ou mean ± sd ; P50 [P25 ; P75]** |
| Chronic renal failure requiring dialysis | 7 (3.6) | 0 (0.0) | 2 (3.8) | 0 (0.0) | 5 (11.9) | 0 (0.0) |
| **Smoking habits (yes)** | 66 (37.7) | 28 (40.6) | 15 (31.9) | 9 (45.0) | 14 (38.9) | 0 (0.0) |
| **Alcohol abuse (yes)** | 44 (25.1) | 24 (34.8) | 7 (14.9) | 7 (35.0) | 6 (16.7) | 0 (0.0) |
| **Glasgow coma score** | 6 [4 ; 8] (3 - 15) | 6 [3 ; 8] | 6 [4 ; 8] | 8 [5 ; 12] | 7 [4 ; 8] | 6 [4 ; 12] |
| **SAPS II** | 49 [41 ; 58] (15 - 92) | 47 [43 ; 56] | 52 [43 ; 63] | 47 [32 ; 62] | 47 [38 ; 53] | 50 [30 ; 57] |
| **Initial mean arterial pressure (mmHg)** | 93 [80 ; 107] | 92 [80 ; 107] | 87 [79 ; 108] | 97 [89 ; 109] | 93 [80 ; 106] | 95 [90 ; 106] |
| **Initial heart rate** | 81 ± 21 | 84 ± 22 | 74 ± 21 | 89 ± 21 | 78 ± 19 | 95 ± 8 |
| **Initial temperature (°C)** | 36.5 [35.8 ; 37.0] | 36.6 [35.8 ; 37.0] | 36.3 [35.5 ; 36.8] | 36.9 [36.1 ; 37.3] | 36.4 [35.7 ; 37.0] | 36.5 [36.0 ; 37.2] |
| **Face trauma** | 45 (22.7) | 44 (55.7) | 1 (1.9) | 0 (0.0) | 0 (0.0) | 0 (0.0) |
| **Chest trauma** | 28 (14.1) | 28 (35.4) | 0 (0.0) | 0 (0.0) | 0 (0.0) | 0 (0.0) |
| **No immunodepressin** | 188 (95.9) | 76 (98.7) | 47 (88.7) | 20 (100.0) | 41 (97.6) | 4 (100.0) |
| **HIV infection** | 1 (0.5) | 0 (0.0) | 1 (1.9) | 0 (0.0) | 0 (0.0) | 0 (0.0) |
| **Long term corticosteroid therapy** | 3 (1.5) | 0 (0.0) | 2 (3.8) | 0 (0.0) | 1 (2.4) | 0 (0.0) |
| **Hematological malignancy** | 0 (0.0) | 0 (0.0) | 0 (0.0) | 0 (0.0) | 0 (0.0) | 0 (0.0) |
| **Cancer** | 4 (2.0) | 1 (1.3) | 3 (5.7) | 0 (0.0) | 0 (0.0) | 0 (0.0) |
| **Immunosuppressive therapy** | 1 (0.5) | 0 (0.0) | 1 (1.9) | 0 (0.0) | 0 (0.0) | 0 (0.0) |
| **Feeding tube** | | | | | | |
| None | 2 (1.6) | 1 (1.9) | 1 (2.9) | 0 (0.0) | 0 (0.0) | 0 (0.0) |
| Oro-gastric tube | 65 (50.4) | 20 (38.5) | 20 (58.8) | 6 (42.9) | 17 (65.4) | 2 (66.7) |
| Naso-gastric tube | 62 (48.1) | 31 (59.6) | 13 (38.2) | 8 (57.1) | 9 (34.6) | 1 (33.3) |
| Intracranial pressure monitoring; n (%) | | | | | | |

| Variable | Brain-injured patients N = 198 | Type of injury | | | | |
|---|---|---|---|---|---|---|
| | | Trauma N = 79 | Subarachnoid hemmorhage N = 53 | Ischemic stroke N = 20 | hematoma N = 42 | Other N = 4 |
| | n (%) or mean ± sd ; P50 [P25 ; P75] | n (%) or mean ± sd ; P50 [P25 ; P75] | n (%) or mean ± sd ; P50 [P25 ; P75] | n (%) or m ean± sd ; P50 [P25 ; P75] | n (%) ou mean ± sd ; P50 [P25 ; P75] ; | n (%) ou mean ± sd ; P50 [P25 ; P75] |
| None | 21 (10.8) | 8 (10.4) | 3 (5.7) | 4 (22.2) | 4 (9.5) | 2 (50.0) |
| Parenchymal monitoring | 95 (49.0) | 63 (81.8) | 7 (13.2) | 9 (50.0) | 16 (38.1) | 0 (0.0) |
| External ventricular drain | 78 (40.2) | 6 (7.8) | 43 (81.1) | 5 (27.8) | 22 (52.4) | 2 (50.0) |
| **Previous infection** | 27 (13.6) | 10 (12.7) | 10 (18.9) | 4 (20.0) | 2 (4.8) | 1 (25.0) |
| **Previous antibiotic therapy** | | | | | | |
| None | 162 (82.7) | 65 (83.3) | 41 (78.8) | 15 (75.0) | 38 (90.5) | 3 (75.0) |
| Beta lactamins | 34 (17.3) | 13 (16.7) | 11 (21.2) | 5 (25.0) | 4 (9.5) | 1 (25.0) |
| Glycopeptide | 0 (0.0) | 0 (0.0) | 0 (0.0) | 0 (0.0) | 0 (0.0) | 0 (0.0) |
| Aminoside | 0 (0.0) | 0 (0.0) | 0 (0.0) | 0 (0.0) | 0 (0.0) | 0 (0.0) |
| Other | 0 (0.0) | 0 (0.0) | 0 (0.0) | 0 (0.0) | 0 (0.0) | 0 (0.0) |
| **VAP Microbiology** | | | | | | |
| Gram negative bacteria | 12 (6.1) | 2 (2.5) | 4 (7.5) | 1 (5.0) | 4 (9.5) | 1 (25.0) |
| Group 0 or 1 Enterbacterales | 7 (3.5) | 5 (6.3) | 0 (0.0) | 1 (5.0) | 1 (2.4) | 0 (0.0) |
| Group 2 Enterbacterales | 19 (9.6) | 7 (8.9) | 6 (11.3) | 3 (15.0) | 3 (7.1) | 0 (0.0) |
| Group 3 Enterbacterales | 13 (6.6) | 6 (7.6) | 2 (3.8) | 2 (10.0) | 2 (4.8) | 1 (25.0) |
| Group 5 Enterbacterales | 0 (0.0) | 0 (0.0) | 0 (0.0) | 0 (0.0) | 0 (0.0) | 0 (0.0) |
| Other Gram negative bactieria | 15 (7.6) | 7 (8.9) | 3 (5.7) | 1 (5.0) | 3 (7.1) | 1 (25.0) |
| Gram positive bacteria | 130 (65.7) | 50 (63.3) | 38 (71.7) | 12 (60.0) | 29 (69.0) | 1 (25.0) |
| Other | 2 (1.0) | 2 (2.5) | 0 (0.0) | 0 (0.0) | 0 (0.0) | 0 (0.0) |
| None | 0 (0.0) | 0 (0.0) | 0 (0.0) | 0 (0.0) | 0 (0.0) | 0 (0.0) |
| **Pathogen: *Staphylococcus aureus*** | 122 (61.6) | 48 (60.8) | 37 (69.8) | 10 (50.0) | 27 (64.3) | 0 (0.0) |
| **Pathogen: *E.coli*** | 27 (13.6) | 15 (19.0) | 6 (11.3) | 2 (10.0) | 3 (7.1) | 1 (25.0) |
| **Pathogen: *Enterobacter cloacae*** | 11 (5.6) | 6 (7.6) | 1 (1.9) | 3 (15.0) | 1 (2.4) | 0 (0.0) |

| Variable | Brain-injured patients N = 198 | Type of injury | | | | |
|---|---|---|---|---|---|---|
| | | Trauma N = 79 | Subarachnoid hemmorhage N = 53 | Ischemic stroke N = 20 | hematoma N = 42 | Other N = 4 |
| | n (%) or mean ± sd ; P50 [P25 ; P75] | n (%) or mean ± sd ; P50 [P25 ; P75] | n (%) or mean ± sd ; P50 [P25 ; P75] | n (%) or m ean± sd ; P50 [P25 ; P75] | n (%) ou mean ± sd ; P50 [P25 ; P75] ; | n (%) ou mean ± sd ; P50 [P25 ; P75] |
| **Pathogen: *Citrobacter Koserii*** | 19 (9.6) | 7 (8.9) | 5 (9.4) | 3 (15.0) | 4 (9.5) | 0 (0.0) |
| **Pathogen: *Klebsiella pneumoniae*** | 25 (12.6) | 14 (17.7) | 6 (11.3) | 2 (10.0) | 3 (7.1) | 0 (0.0) |
| **Pathogen: *Enterobacter aerogenes*** | 13 (6.6) | 5 (6.3) | 3 (5.7) | 2 (10.0) | 3 (7.1) | 0 (0.0) |
| **Pathogen: *Pseudomonas aeruginosa*** | 18 (9.1) | 5 (6.3) | 5 (9.4) | 2 (10.0) | 5 (11.9) | 1 (25.0) |
| **Pathogène : other** | 111 (56.1) | 44 (55.7) | 29 (54.7) | 11 (55.0) | 23 (54.8) | 4 (100.0) |
| **Staphylococcus aureus during VAP** | | | | | | |
| Single pathogen | 33 (27.0) | 13 (27.1) | 12 (32.4) | 0 (0.0) | 8 (29.6) | - |
| Multiple pathogens | 89 (73.0) | 35 (72.9) | 25 (67.6) | 10 (100.0) | 19 (70.4) | - |
| **ICU length of stay (days)** | 27 [15 ; 40] | 26 [16 ; 44] | 27 [15 ; 39] | 22 [13 ; 34] | 30 [16 ; 41] | 36 [21 ; 54] |
| **Length of invasive mechanical ventilation (days)** | 21 [13 ; 32] | 22 [13 ; 31] | 19 [12 ; 32] | 17 [12 ; 25] | 22 [14 ; 38] | 28 [18 ; 39] |
| **Secondary ischemic stroke** | 24 (12.3) | 1 (1.3) | 19 (36.5) | 0 (0.0) | 3 (7.3) | 1 (33.3) |
| **Septic shock** | 62 (32.0) | 24 (30.4) | 19 (38.0) | 4 (20.0) | 13 (31.7) | 2 (50.0) |
| **Mortality at D28 or D90** | 77 (42.8) | 30 (41.7) | 24 (50.0) | 7 (38.9) | 15 (38.5) | 1 (33.3) |
| **CPIS score at VAP time** | 6 [4 ; 7] | 6 [4 ; 7] | 6 [4 ; 7] | 6 [4 ; 6] | 4 [3 ; 6] | 6 [5 ; 7] |
| **Delay of VAP (days)** | 6 [4 ; 8] | 6 [4 ; 8] | 5 [4 ; 7] | 7 [6 ; 8] | 6 [4 ; 9] | 9 [5 ; 14] |
| **Initial glycemia (mmol/L)** | 7.63 [6.50 ; 9.70] | 7.15 [6.00 ; 9.10] | 7.85 [6.90 ; 10.20] | 7.70 [6.97 ; 9.63] | 7.80 [6.60 ; 9.80] | 7.50 [5.50 ; 8.20] |
| **Albumin (g/L)** | 36.1 ± 5.9 | 33.8 ± 5.2 | 37.6 ± 6.0 | 39.4 ± 7.7 | 38.3 ± 4.8 | 30.0 ± NA |
| **Leucocytes (G/L)** | 13.20 [10.40 ; 17.70] | 14.50 [11.80 ; 20.40] | 14.30 [10.30 ; 17.70] | 12.30 [10.40 ; 14.60] (7.60 - 22.90) | 11.90 [9.35 ; 14.40] (5.60 - 19.60) | 9.00 [5.95 ; 14.00] (4.70 - 17.20) |

| Variable | Brain-injured patients N = 198 | Type of injury | | | | |
|---|---|---|---|---|---|---|
| | | Trauma N = 79 | Subarachnoid hemmorhage N = 53 | Ischemic stroke N = 20 | hematoma N = 42 | Other N = 4 |
| | n (%) or mean ± sd ; P50 [P25 ; P75] | n (%) or mean ± sd ; P50 [P25 ; P75] | n (%) or mean ± sd ; P50 [P25 ; P75] | n (%) or m ean± sd ; P50 [P25 ; P75] | n (%) ou mean ± sd ; P50 [P25 ; P75] ; | n (%) ou mean ± sd ; P50 [P25 ; P75] |
| **CPK (UI/L)** | 336 [152 ; 757] | 485 [283 ; 1,015] | 152 [97 ; 347] | 261 [148 ; 662] | 208 [119 ; 681] | 595 [101 ; 1,088] |
| **Bilirubin (Umol/L)** | 9 [7 ; 13] | 9 [6 ; 12] | 9 [7 ; 12] | 10 [6 ; 13] | 10 [6 ; 16] | 14 [12 ; 15] |
| **Natremia (mmol/L)** | 138 [136 ; 141] | 138 [136 ; 140] | 138 [135 ; 142] (127 - 157) | 139 [136 ; 143] (130 - 148) | 139 [136 ; 140] (126 - 160) | 136 [135 ; 139] (135 - 142) |
| **Créatinin (umol/L)** | 72 [57 ; 94] | 78 [63 ; 102] (34 - 331) | 63 [53 ; 79] (18 - 753) | 80 [57 ; 168] (33 - 227) | 73 [57 ; 109] (33 - 991) | 41 [36 ; 59] (31 - 76) |
| **Blood urea nitrogen (mmol/L)** | 4.75 [3.70 ; 6.16] | 4.60 [3.30 ; 6.20] | 4.80 [4.00 ; 5.80] | 5.20 [3.90 ; 6.90] | 4.70 [4.18 ; 7.80] | 4.85 [4.20 ; 8.25] |
| **Blood transfusion** | | | | | | |
| None | 69 (52.7) | 17 (34.7) | 26 (70.3) | 10 (76.9) | 14 (50.0) | 2 (50.0) |
| <= 3 blood bag | 45 (34.4) | 20 (40.8) | 10 (27.0) | 3 (23.1) | 10 (35.7) | 2 (50.0) |
| > 3 | 17 (13.0) | 12 (24.5) | 1 (2.7) | 0 (0.0) | 4 (14.3) | 0 (0.0) |
| **Proton pump inhibitors** | 32 (50.0) | 7 (43.8) | 8 (44.4) | 3 (50.0) | 14 (63.6) | 0 (0.0) |
| **Corticosteroid therapy** | 53 (57.6) | 19 (57.6) | 18 (66.7) | 2 (25.0) | 12 (57.1) | 2 (66.7) |
| **Insulin** | 82 (100.0) | 20 (100.0) | 27 (100.0) | 9 (100.0) | 23 (100.0) | 3 (100.0) |
| **Propofol** | 179 (92.3) | 72 (92.3) | 49 (94.2) | 17 (89.5) | 38 (92.7) | 3 (75.0) |
| **Midazolam** | 146 (75.3) | 65 (83.3) | 41 (78.8) | 10 (52.6) | 28 (68.3) | 2 (50.0) |
| **Sufentanyl** | 174 (89.7) | 73 (93.6) | 49 (94.2) | 14 (73.7) | 35 (85.4) | 3 (75.0) |
| **Orhter** | 127 (65.5) | 60 (76.9) | 37 (71.2) | 8 (42.1) | 20 (48.8) | 2 (50.0) |
| **Thoraci surgery** | 9 (4.5) | 6 (7.6) | 1 (1.9) | 0 (0.0) | 2 (4.8) | 0 (0.0) |
| **Neurosurgery** | 147 (74.2) | 49 (62.0) | 44 (83.0) | 19 (95.0) | 32 (76.2) | 3 (75.0) |
| **Orthopédic surgery** | 17 (8.6) | 17 (21.5) | 0 (0.0) | 0 (0.0) | 0 (0.0) | 0 (0.0) |

| Variable | Brain-injured patients N = 198 | Type of injury | | | | |
|---|---|---|---|---|---|---|
| | | Trauma N = 79 | Subarachnoid hemmorhage N = 53 | Ischemic stroke N = 20 | hematoma N = 42 | Other N = 4 |
| | n (%) or mean ± sd ; P50 [P25 ; P75] | n (%) or mean ± sd ; P50 [P25 ; P75] | n (%) or mean ± sd ; P50 [P25 ; P75] | n (%) or m ean± sd ; P50 [P25 ; P75] | n (%) ou mean ± sd ; P50 [P25 ; P75] ; | n (%) ou mean ± sd ; P50 [P25 ; P75] |
| **Other surgery** | 25 (12.6) | 11 (13.9) | 8 (15.1) | 0 (0.0) | 6 (14.3) | 0 (0.0) |
| **ECMO** | 2 (1.0) | 2 (2.5) | 0 (0.0) | 0 (0.0) | 0 (0.0) | 0 (0.0) |
| **Intermitent dialysis** | 9 (4.5) | 2 (2.5) | 4 (7.5) | 0 (0.0) | 3 (7.1) | 0 (0.0) |
| **Continuous veinoveinous dialysis** | 19 (9.6) | 10 (12.7) | 4 (7.5) | 0 (0.0) | 5 (11.9) | 0 (0.0) |
| **Delay to initial tracheal intubation** | 1 [1 ; 1] (1 - 72) | 1 [1 ; 1] (1 - 72) | 1 [1 ; 1] (1 - 24) | 1 [1 ; 24] (1 - 48) | 1 [1 ; 1] (1 - 72) | 2 [1 ; 10] (1 - 17) |
| **Length of invasive mechanical ventilation** | 21 [13 ; 31] | 22 [13 ; 31] | 19 [13 ; 32] | 17 [12 ; 25] | 22 [14 ; 35] | 25 [18 ; 33] |
| **Targeted temperature contol** | 59 (29.9) | 32 (41.0) | 14 (26.4) | 4 (20.0) | 8 (19.0) | 1 (25.0) |
| **Prone position** | | | | | | |
| None | 174 (87.9) | 64 (81.0) | 48 (90.6) | 19 (95.0) | 39 (92.9) | 4 (100.0) |
| <= 2 | 14 (7.1) | 9 (11.4) | 2 (3.8) | 1 (5.0) | 2 (4.8) | 0 (0.0) |
| > 2 | 10 (5.1) | 6 (7.6) | 3 (5.7) | 0 (0.0) | 1 (2.4) | 0 (0.0) |

*Nombre de données disponibles

BMI: body mass index; COPD: chronic obstructive pulmonary disease; ECMO: extra-corporeal membrane oxygenation; VAP: ventilator-associated pneumonia; SAPS II: simplified acute physiology score.

**Supplementary table 2**: pathogens associated with Staphylococcus aureus during VAP

| Combination of pathogens | others | n |
|---|---|---|
| *Staphylococcus aureus* + other | *C. .freundi, A.baumanii* | 1 |
| | *C.koseri* | 1 |
| | *C. freundii, H. influenzae, Pantoea agglmomerans* | 1 |
| | *H. alvei* | 1 |
| | *H. alvei, G. vaginalis* | 1 |
| | *H. alvei, S. pneumoniae,* | 1 |
| | *H. influenzae* | 23 |
| | *H. influenzae, S. pneumoniae* | 2 |
| | *H. influenzae, S. anginosus* | 1 |
| | *E.aerogenes* | 1 |
| | *K. ocytoca, P. vulgaris* | 1 |
| | *K. oxytoca* | 5 |
| | *K. oxytoca; H. influenzae* | 1 |
| | *P. mirabilis* | 1 |
| | *S. pneumoniae* | 1 |
| | *S. anginosus* | 1 |
| | *S. anginosus, S. constellatus* | 1 |
| | *S. constellatus* | 3 |
| | *S. marcescens* | 1 |
| | *S. pneumoniae* | 2 |
| | *S. urealytica, S. marcesens* | 1 |
| | *S. marcescens, S. anginosus* | 1 |
| | *S. marcescens* | 1 |
| | *S. anginosus* | 1 |
| | *S. anginosus, P. mirabilis* | 1 |
| | *S. constellatus, S. oralis, N. macacae* | 1 |
| *Staphylococcus aureus + E.coli* + other | *H. influenzae* | 4 |

| **Combination of pathogens** | **others** | **n** |
|---|---|---|
| | *H.influenzae, S. anginosus* | 1 |
| | *K. oxytoca, A. baumanii* | 1 |
| | *M.catarrhalis* | 1 |
| | *M. morgagnii* | 1 |
| | *S. agalactiae* | 1 |
| | *S. marcescens, S. maltophilia, A. baumanii* | 1 |
| *Staphylococcus aureus + Klebsiella pneumoniae* + other | *A. baumanii* | 1 |
| | *E. faecalis, S. marcescens* | 1 |
| | *H. influenzae* | 1 |
| | *H. influenzae, C. freundii,* | 1 |
| | *H. influenzae* | 1 |
| | *M morgangii* | 1 |
| | *P. mirabilis* | 1 |
| | *S. pneumoniae, H. influenzae* | 1 |
| *Staphylococcus aureus + Enterobacter aerogenes* + other | *A. baumanii* | 1 |
| | *M. morgagnii, P. vulgaris* | 1 |
| | *S. pneumoniae, Bacillus cereus* | 1 |
| | *S. neumoniae* | 1 |
| | *S. anginosus, S. oralis, E. faecalis,* | 1 |
| | *H.influenzae* | 2 |
| | *K oxytoca* | 1 |
| | *S. pneumoniae* | 1 |
| *Staphylococcus aureus + Citrobacter Koserii* + other | *H. influenzae* | 2 |
| | *M. catarrhalis* | 1 |
| *Staphylococcus aureus + E.coli + Enterobacter aerogenes* + other | *H. influenzae, S. maltophilia* | 1 |
| | *S. marcescens, A. baumanii* | 1 |
| *Staphylococcus aureus + Citrobacter Koseri + Enterobacter aerogenes* + other | *H. influenzae* | 1 |

| **Combination of pathogens** | **others** | **n** |
|---|---|---|
| *Staphylococccus aureus + E.coli + Citrobacter Koserii* + Other | *S. anginosus* | 1 |
| *Staphylococcus aureus + E.coli + Klebsiella pneumoniae + Pseudomonas aeruginosa* + other | *H. influenzae* | 1 |
| *Staphylococcus aureus + Klebsiella pneumoniae + Enterobacter aerogenes* + Other | *H. influenzae* | 1 |

**Supplementary table 3**: initial characteristics of brain-injured patients

| Variable | Brain-injured patients N = 198 | Type of injury | | | | |
|---|---|---|---|---|---|---|
| | | Trauma N = 79 | Subarachnoid hemorrhage N = 53 | Ischemic stroke N = 20 | hematoma N = 42 | Other N = 4 |
| | n (%) or mean ± sd; P50 [P25; P75] | n (%) or mean ± sd; P50 [P25; P75] | n (%) or mean ± sd; P50 [P25; P75] | n (%) or mean± sd; P50 [P25; P75] | n (%) or mean ± sd; P50 [P25; P75] | n (%) or mean ± sd; P50 [P25; P75] |
| **Sex** | | | | | | |
| Male | 134 (67.7) | 67 (84.8) | 27 (50.9) | 13 (65.0) | 25 (59.5) | 2 (50.0) |
| Female | 64 (32.3) | 12 (15.2) | 26 (49.1) | 7 (35.0) | 17 (40.5) | 2 (50.0) |
| **Age (years)** | 52 [36 ; 63] | 44 [27 ; 59] | 56 [49 ; 67] | 47 [35 ; 59] | 54 [41 ; 63] | 58 [53 ; 69] |
| **BMI (kg/m2)** | 24.00 [21.17 ; 27.80] (13.50 - 45.80) | 22.90 [20.30 ; 25.50] (13.50 - 39.50) | 24.69 [21.30 ; 29.38] (16.98 - 45.80) | 26.60 [22.90 ; 28.10] (18.52 - 37.90) | 24.84 [21.46 ; 28.34] (16.00 - 42.24) | 23.23 [21.10 ; 27.50] (21.10 - 27.50) |
| **No cardiac or vascular disease** | 144 (73.5) | 64 (83.1) | 35 (66.0) | 16 (80.0) | 27 (64.3) | 2 (50.0) |
| **Chronic heart failure** | | | | | | |
| No | 192 (98.0) | 75 (97.4) | 53 (100.0) | 19 (95.0) | 41 (97.6) | 4 (100.0) |
| yes | 4 (2.0) | 2 (2.6) | 0 (0.0) | 1 (5.0) | 1 (2.4) | 0 (0.0) |
| **Arteritis** | 2 (1.0) | 1 (1.3) | 1 (1.9) | 0 (0.0) | 0 (0.0) | 0 (0.0) |
| **Diabetes mellitus** | | | | | | |
| No | 175 (89.3) | 70 (90.9) | 45 (84.9) | 19 (95.0) | 38 (90.5) | 3 (75.0) |
| uncomplicated | 13 (6.6) | 4 (5.2) | 6 (11.3) | 1 (5.0) | 1 (2.4) | 1 (25.0) |
| Complicated | 8 (4.1) | 3 (3.9) | 2 (3.8) | 0 (0.0) | 3 (7.1) | 0 (0.0) |
| **Hypertension** | 29 (14.8) | 4 (5.2) | 10 (18.9) | 2 (10.0) | 12 (28.6) | 1 (25.0) |
| **Atrial fibrillation** | 3 (1.5) | 1 (1.3) | 1 (1.9) | 0 (0.0) | 1 (2.4) | 0 (0.0) |

| Variable | Brain-injured patients N = 198 | Type of injury | | | | |
|---|---|---|---|---|---|---|
| | | Trauma N = 79 | Subarachnoid hemorrhage N = 53 | Ischemic stroke N = 20 | hematoma N = 42 | Other N = 4 |
| | n (%) or mean ± sd; P50 [P25; P75] | n (%) or mean ± sd; P50 [P25; P75] | n (%) or mean ± sd; P50 [P25; P75] | n (%) or mean± sd; P50 [P25; P75] | n (%) or mean ± sd; P50 [P25; P75] | n (%) or mean ± sd; P50 [P25; P75] |
| **COPD** | 5 (2.6) | 3 (3.9) | 1 (1.9) | 0 (0.0) | 1 (2.4) | 0 (0.0) |
| **Chronic respiratory failure** | 1 (0.5) | 0 (0.0) | 0 (0.0) | 0 (0.0) | 1 (2.4) | 0 (0.0) |
| **Long-term oxygen** | 0 (0.0) | 0 (0.0) | 0 (0.0) | 0 (0.0) | 0 (0.0) | 0 (0.0) |
| **Cirrhosis** | 1 (0.5) | 1 (1.3) | 0 (0.0) | 0 (0.0) | 0 (0.0) | 0 (0.0) |
| Chronic renal failure | 5 (2.6) | 1 (1.3) | 0 (0.0) | 4 (20.0) | 0 (0.0) | 0 (0.0) |
| Chronic renal failure requiring dialysis | 7 (3.6) | 0 (0.0) | 2 (3.8) | 0 (0.0) | 5 (11.9) | 0 (0.0) |
| **Smoking habits (yes)** | 66 (37.7) | 28 (40.6) | 15 (31.9) | 9 (45.0) | 14 (38.9) | 0 (0.0) |
| **Alcohol abuse (yes)** | 44 (25.1) | 24 (34.8) | 7 (14.9) | 7 (35.0) | 6 (16.7) | 0 (0.0) |
| **Glasgow coma score** | 6 [4 ; 8] (3 - 15) | 6 [3 ; 8] | 6 [4 ; 8] | 8 [5 ; 12] | 7 [4 ; 8] | 6 [4 ; 12] |
| **SAPS II** | 49 [41 ; 58] (15 - 92) | 47 [43 ; 56] | 52 [43 ; 63] | 47 [32 ; 62] | 47 [38 ; 53] | 50 [30 ; 57] |
| **Initial mean arterial pressure (mmHg)** | 93 [80 ; 107] | 92 [80 ; 107] | 87 [79 ; 108] | 97 [89 ; 109] | 93 [80 ; 106] | 95 [90 ; 106] |
| **Initial heart rate** | 81 ± 21 | 84 ± 22 | 74 ± 21 | 89 ± 21 | 78 ± 19 | 95 ± 8 |
| **Initial temperature (°C)** | 36.5 [35.8 ; 37.0] | 36.6 [35.8 ; 37.0] | 36.3 [35.5 ; 36.8] | 36.9 [36.1 ; 37.3] | 36.4 [35.7 ; 37.0] | 36.5 [36.0 ; 37.2] |
| **Face trauma** | 45 (22.7) | 44 (55.7) | 1 (1.9) | 0 (0.0) | 0 (0.0) | 0 (0.0) |
| **Chest trauma** | 28 (14.1) | 28 (35.4) | 0 (0.0) | 0 (0.0) | 0 (0.0) | 0 (0.0) |
| **No immunodepressin** | 188 (95.9) | 76 (98.7) | 47 (88.7) | 20 (100.0) | 41 (97.6) | 4 (100.0) |
| **HIV infection** | 1 (0.5) | 0 (0.0) | 1 (1.9) | 0 (0.0) | 0 (0.0) | 0 (0.0) |
| **Long term corticosteroid therapy** | 3 (1.5) | 0 (0.0) | 2 (3.8) | 0 (0.0) | 1 (2.4) | 0 (0.0) |

| Variable | Brain-injured patients N = 198 | Type of injury | | | | |
|---|---|---|---|---|---|---|
| | | Trauma N = 79 | Subarachnoid hemorrhage N = 53 | Ischemic stroke N = 20 | hematoma N = 42 | Other N = 4 |
| | n (%) or mean ± sd; P50 [P25; P75] | n (%) or mean ± sd; P50 [P25; P75] | n (%) or mean ± sd; P50 [P25; P75] | n (%) or mean± sd; P50 [P25; P75] | n (%) or mean ± sd; P50 [P25; P75] | n (%) or mean ± sd; P50 [P25; P75] |
| **Hematological malignancy** | 0 (0.0) | 0 (0.0) | 0 (0.0) | 0 (0.0) | 0 (0.0) | 0 (0.0) |
| **Cancer** | 4 (2.0) | 1 (1.3) | 3 (5.7) | 0 (0.0) | 0 (0.0) | 0 (0.0) |
| **Immunosuppressive therapy** | 1 (0.5) | 0 (0.0) | 1 (1.9) | 0 (0.0) | 0 (0.0) | 0 (0.0) |
| **Feeding tube** | | | | | | |
| None | 2 (1.6) | 1 (1.9) | 1 (2.9) | 0 (0.0) | 0 (0.0) | 0 (0.0) |
| Oro-gastric tube | 65 (50.4) | 20 (38.5) | 20 (58.8) | 6 (42.9) | 17 (65.4) | 2 (66.7) |
| Naso-gastric tube | 62 (48.1) | 31 (59.6) | 13 (38.2) | 8 (57.1) | 9 (34.6) | 1 (33.3) |
| Intracranial pressure monitoring; n (%) | | | | | | |
| None | 21 (10.8) | 8 (10.4) | 3 (5.7) | 4 (22.2) | 4 (9.5) | 2 (50.0) |
| Parenchymal monitoring | 95 (49.0) | 63 (81.8) | 7 (13.2) | 9 (50.0) | 16 (38.1) | 0 (0.0) |
| External ventricular drain | 78 (40.2) | 6 (7.8) | 43 (81.1) | 5 (27.8) | 22 (52.4) | 2 (50.0) |
| **Previous infection** | 27 (13.6) | 10 (12.7) | 10 (18.9) | 4 (20.0) | 2 (4.8) | 1 (25.0) |
| **Previous antibiotic therapy** | | | | | | |
| None | 162 (82.7) | 65 (83.3) | 41 (78.8) | 15 (75.0) | 38 (90.5) | 3 (75.0) |
| Beta lactamins | 34 (17.3) | 13 (16.7) | 11 (21.2) | 5 (25.0) | 4 (9.5) | 1 (25.0) |
| Glycopeptide | 0 (0.0) | 0 (0.0) | 0 (0.0) | 0 (0.0) | 0 (0.0) | 0 (0.0) |
| Aminoside | 0 (0.0) | 0 (0.0) | 0 (0.0) | 0 (0.0) | 0 (0.0) | 0 (0.0) |
| Other | 0 (0.0) | 0 (0.0) | 0 (0.0) | 0 (0.0) | 0 (0.0) | 0 (0.0) |
| **VAP Microbiology** | | | | | | |
| Gram negative bacteria | 12 (6.1) | 2 (2.5) | 4 (7.5) | 1 (5.0) | 4 (9.5) | 1 (25.0) |
| Group 0 or 1 Enterbacterales | 7 (3.5) | 5 (6.3) | 0 (0.0) | 1 (5.0) | 1 (2.4) | 0 (0.0) |

| Variable | Brain-injured patients N = 198 | Type of injury | | | | |
|---|---|---|---|---|---|---|
| | | Trauma N = 79 | Subarachnoid hemorrhage N = 53 | Ischemic stroke N = 20 | hematoma N = 42 | Other N = 4 |
| | n (%) or mean ± sd; P50 [P25; P75] | n (%) or mean ± sd; P50 [P25; P75] | n (%) or mean ± sd; P50 [P25; P75] | n (%) or mean± sd; P50 [P25; P75] | n (%) or mean ± sd; P50 [P25; P75] | n (%) or mean ± sd; P50 [P25; P75] |
| Group 2 Enterbacterales | 19 (9.6) | 7 (8.9) | 6 (11.3) | 3 (15.0) | 3 (7.1) | 0 (0.0) |
| Group 3 Enterbacterales | 13 (6.6) | 6 (7.6) | 2 (3.8) | 2 (10.0) | 2 (4.8) | 1 (25.0) |
| Group 5 Enterbacterales | 0 (0.0) | 0 (0.0) | 0 (0.0) | 0 (0.0) | 0 (0.0) | 0 (0.0) |
| Other Gram negative bactieria | 15 (7.6) | 7 (8.9) | 3 (5.7) | 1 (5.0) | 3 (7.1) | 1 (25.0) |
| Gram positive bacteria | 130 (65.7) | 50 (63.3) | 38 (71.7) | 12 (60.0) | 29 (69.0) | 1 (25.0) |
| Other | 2 (1.0) | 2 (2.5) | 0 (0.0) | 0 (0.0) | 0 (0.0) | 0 (0.0) |
| None | 0 (0.0) | 0 (0.0) | 0 (0.0) | 0 (0.0) | 0 (0.0) | 0 (0.0) |
| **Pathogen: *Staphylococcus aureus*** | 122 (61.6) | 48 (60.8) | 37 (69.8) | 10 (50.0) | 27 (64.3) | 0 (0.0) |
| **Pathogen: *E. coli*** | 27 (13.6) | 15 (19.0) | 6 (11.3) | 2 (10.0) | 3 (7.1) | 1 (25.0) |
| **Pathogen: *Enterobacter cloacae*** | 11 (5.6) | 6 (7.6) | 1 (1.9) | 3 (15.0) | 1 (2.4) | 0 (0.0) |
| **Pathogen: *Citrobacter Koserii*** | 19 (9.6) | 7 (8.9) | 5 (9.4) | 3 (15.0) | 4 (9.5) | 0 (0.0) |
| **Pathogen: *Klebsiella pneumoniae*** | 25 (12.6) | 14 (17.7) | 6 (11.3) | 2 (10.0) | 3 (7.1) | 0 (0.0) |
| **Pathogen: *Enterobacter aerogenes*** | 13 (6.6) | 5 (6.3) | 3 (5.7) | 2 (10.0) | 3 (7.1) | 0 (0.0) |
| **Pathogen: *Pseudomonas aeruginosa*** | 18 (9.1) | 5 (6.3) | 5 (9.4) | 2 (10.0) | 5 (11.9) | 1 (25.0) |
| **Pathogène : other** | 111 (56.1) | 44 (55.7) | 29 (54.7) | 11 (55.0) | 23 (54.8) | 4 (100.0) |
| **Staphylococcus aureus during VAP** | | | | | | |
| Single pathogen | 33 (27.0) | 13 (27.1) | 12 (32.4) | 0 (0.0) | 8 (29.6) | - |
| Multiple pathogens | 89 (73.0) | 35 (72.9) | 25 (67.6) | 10 (100.0) | 19 (70.4) | - |
| **ICU length of stay (days)** | 27 [15; 40] | 26 [16; 44] | 27 [15; 39] | 22 [13; 34] | 30 [16; 41] | 36 [21; 54] |
| **Length of invasive mechanical ventilation (days)** | 21 [13; 32] | 22 [13; 31] | 19 [12; 32] | 17 [12; 25] | 22 [14; 38] | 28 [18; 39] |
| **Secondary ischemic stroke** | 24 (12.3) | 1 (1.3) | 19 (36.5) | 0 (0.0) | 3 (7.3) | 1 (33.3) |

| Variable | Brain-injured patients N = 198 | Type of injury | | | | |
|---|---|---|---|---|---|---|
| | | Trauma N = 79 | Subarachnoid hemorrhage N = 53 | Ischemic stroke N = 20 | hematoma N = 42 | Other N = 4 |
| | n (%) or mean ± sd; P50 [P25; P75] | n (%) or mean ± sd; P50 [P25; P75] | n (%) or mean ± sd; P50 [P25; P75] | n (%) or mean± sd; P50 [P25; P75] | n (%) or mean ± sd; P50 [P25; P75] | n (%) or mean ± sd; P50 [P25; P75] |
| **Septic shock** | 62 (32.0) | 24 (30.4) | 19 (38.0) | 4 (20.0) | 13 (31.7) | 2 (50.0) |
| **Mortality at D28 or D90** | 77 (42.8) | 30 (41.7) | 24 (50.0) | 7 (38.9) | 15 (38.5) | 1 (33.3) |
| **CPIS score at VAP time** | 6 [4; 7] | 6 [4; 7] | 6 [4; 7] | 6 [4; 6] | 4 [3; 6] | 6 [5; 7] |
| **Delay of VAP (days)** | 6 [4; 8] | 6 [4; 8] | 5 [4; 7] | 7 [6; 8] | 6 [4; 9] | 9 [5; 14] |
| **Initial glycemia (mmol/L)** | 7.63 [6.50; 9.70] | 7.15 [6.00; 9.10] | 7.85 [6.90; 10.20] | 7.70 [6.97; 9.63] | 7.80 [6.60; 9.80] | 7.50 [5.50; 8.20] |
| **Albumin (g/L)** | 36.1 ± 5.9 | 33.8 ± 5.2 | 37.6 ± 6.0 | 39.4 ± 7.7 | 38.3 ± 4.8 | 30.0 ± NA |
| **Leucocytes (G/L)** | 13.20 [10.40; 17.70] | 14.50 [11.80; 20.40] | 14.30 [10.30; 17.70] | 12.30 [10.40; 14.60] (7.60 - 22.90) | 11.90 [9.35; 14.40] (5.60 - 19.60) | 9.00 [5.95; 14.00] (4.70 - 17.20) |
| **CPK (UI/L)** | 336 [152; 757] | 485 [283; 1,015] | 152 [97; 347] | 261 [148; 662] | 208 [119; 681] | 595 [101; 1,088] |
| **Bilirubin (Umol/L)** | 9 [7; 13] | 9 [6; 12] | 9 [7; 12] | 10 [6; 13] | 10 [6; 16] | 14 [12; 15] |
| **Natremia (mmol/L)** | 138 [136; 141] | 138 [136; 140] | 138 [135; 142] (127 - 157) | 139 [136; 143] (130 - 148) | 139 [136; 140] (126 - 160) | 136 [135; 139] (135 - 142) |
| **Créatinin (umol/L)** | 72 [57; 94] | 78 [63; 102] (34 - 331) | 63 [53; 79] (18 - 753) | 80 [57; 168] (33 - 227) | 73 [57; 109] (33 - 991) | 41 [36; 59] (31 - 76) |
| **Blood urea nitrogen (mmol/L)** | 4.75 [3.70; 6.16] | 4.60 [3.3; 6.2] | 4.80 [4.0; 5.8] | 5.20 [3.9; 6.9] | 4.70 [4.2; 7.8] | 4.85 [4.2; 8.3] |
| **Blood transfusion** | | | | | | |
| None | 69 (52.7) | 17 (34.7) | 26 (70.3) | 10 (76.9) | 14 (50.0) | 2 (50.0) |
| <= 3 blood bag | 45 (34.4) | 20 (40.8) | 10 (27.0) | 3 (23.1) | 10 (35.7) | 2 (50.0) |
| > 3 | 17 (13.0) | 12 (24.5) | 1 (2.7) | 0 (0.0) | 4 (14.3) | 0 (0.0) |

| Variable | Brain-injured patients N = 198 | Type of injury: Trauma N = 79 | Subarachnoid hemorrhage N = 53 | Ischemic stroke N = 20 | hematoma N = 42 | Other N = 4 |
|---|---|---|---|---|---|---|
| | n (%) or mean ± sd; P50 [P25; P75] | n (%) or mean ± sd; P50 [P25; P75] | n (%) or mean ± sd; P50 [P25; P75] | n (%) or mean± sd; P50 [P25; P75] | n (%) or mean ± sd; P50 [P25; P75] | n (%) or mean ± sd; P50 [P25; P75] |
| **Proton pump inhibitors** | 32 (50.0) | 7 (43.8) | 8 (44.4) | 3 (50.0) | 14 (63.6) | 0 (0.0) |
| **Corticosteroid therapy** | 53 (57.6) | 19 (57.6) | 18 (66.7) | 2 (25.0) | 12 (57.1) | 2 (66.7) |
| **Insulin** | 82 (100.0) | 20 (100.0) | 27 (100.0) | 9 (100.0) | 23 (100.0) | 3 (100.0) |
| **Propofol** | 179 (92.3) | 72 (92.3) | 49 (94.2) | 17 (89.5) | 38 (92.7) | 3 (75.0) |
| **Midazolam** | 146 (75.3) | 65 (83.3) | 41 (78.8) | 10 (52.6) | 28 (68.3) | 2 (50.0) |
| **Sufentanyl** | 174 (89.7) | 73 (93.6) | 49 (94.2) | 14 (73.7) | 35 (85.4) | 3 (75.0) |
| **Orhter** | 127 (65.5) | 60 (76.9) | 37 (71.2) | 8 (42.1) | 20 (48.8) | 2 (50.0) |
| **Thoraci surgery** | 9 (4.5) | 6 (7.6) | 1 (1.9) | 0 (0.0) | 2 (4.8) | 0 (0.0) |
| **Neurosurgery** | 147 (74.2) | 49 (62.0) | 44 (83.0) | 19 (95.0) | 32 (76.2) | 3 (75.0) |
| **Orthopédic surgery** | 17 (8.6) | 17 (21.5) | 0 (0.0) | 0 (0.0) | 0 (0.0) | 0 (0.0) |
| **Other surgery** | 25 (12.6) | 11 (13.9) | 8 (15.1) | 0 (0.0) | 6 (14.3) | 0 (0.0) |
| **ECMO** | 2 (1.0) | 2 (2.5) | 0 (0.0) | 0 (0.0) | 0 (0.0) | 0 (0.0) |
| **Intermitent dialysis** | 9 (4.5) | 2 (2.5) | 4 (7.5) | 0 (0.0) | 3 (7.1) | 0 (0.0) |
| **Continuous veinoveinous dialysis** | 19 (9.6) | 10 (12.7) | 4 (7.5) | 0 (0.0) | 5 (11.9) | 0 (0.0) |
| **Delay to initial tracheal intubation** | 1 [1; 1] (1 - 72) | 1 [1; 1] (1 - 72) | 1 [1; 1] (1 - 24) | 1 [1; 24] (1 - 48) | 1 [1; 1] (1 - 72) | 2 [1; 10] (1 - 17) |
| **Length of invasive mechanical ventilation** | 21 [13; 31] | 22 [13; 31] | 19 [13; 32] | 17 [12; 25] | 22 [14; 35] | 25 [18; 33] |
| **Targeted temperature contol** | 59 (29.9) | 32 (41.0) | 14 (26.4) | 4 (20.0) | 8 (19.0) | 1 (25.0) |
| **Prone position** | | | | | | |
| None | 174 (87.9) | 64 (81.0) | 48 (90.6) | 19 (95.0) | 39 (92.9) | 4 (100.0) |
| <= 2 | 14 (7.1) | 9 (11.4) | 2 (3.8) | 1 (5.0) | 2 (4.8) | 0 (0.0) |

| Variable | Brain-injured patients N = 198 | Type of injury | | | | |
|---|---|---|---|---|---|---|
| | | Trauma N = 79 | Subarachnoid hemorrhage N = 53 | Ischemic stroke N = 20 | hematoma N = 42 | Other N = 4 |
| | n (%) or mean ± sd; P50 [P25; P75] | n (%) or mean ± sd; P50 [P25; P75] | n (%) or mean ± sd; P50 [P25; P75] | n (%) or mean± sd; P50 [P25; P75] | n (%) or mean ± sd; P50 [P25; P75] | n (%) or mean ± sd; P50 [P25; P75] |
| > 2 | 10 (5.1) | 6 (7.6) | 3 (5.7) | 0 (0.0) | 1 (2.4) | 0 (0.0) |

*Nombre de données disponibles

BMI: body mass index; COPD: chronic obstructive pulmonary disease; ECMO: extra-corporeal membrane oxygenation; VAP: ventilator-associated pneumonia; SAPS II: simplified acute physiology score.

**Supplementary table 4**: backward multivariate analysis in the whole population: percentage of times the variable is selected in the model

| **Variable** | percentage of times the variable is selected in the model (40 réplications) **(%)** |
|---|---|
| Antibioprophylaxis | 100 |
| Previous bta-lactamine | 100 |
| Sub-arachnoid hemorrhage | 100 |
| Age (years) | 100 |
| Arteritis | 100 |
| SAPS II | 100 |
| Thoracic surgery | 100 |
| Length of tracheal intubation (days) | 100 |
| Cirrhosis | 77.5 |
| Glasgow coma score | 57.5 |
| Midazolam | 32.5 |
| Blood urea nitrogen (mmol/L) | 5 |
| Chronic heart failure | 5 |
| Hemorragic stroke | 2.5 |
| Créatinine(umol/L) | 2.5 |
| Face trauma | 2.5 |

**Supplementary table 5:** backward multivariate analysis in the brain-injured population: percentage of times the variable is selected in the model

| **Variable** | percentage of times the variable is selected in the model (40 réplications) **(%)** |
|---|---|
| Arteritis | 100 |
| SAPS II | 100 |
| Sub-arachnoid hemorrhage | 82.5 |
| Chronic respiratory failure | 82.5 |
| Glasgow coma score | 82.5 |
| Creatinin (µmol/l) | 77.5 |
| Cirrhosis | 65 |
| Initial hypothermia | 57.5 |
| Midazolam | 30 |
| Intracranial pressure monitoring | 17.5 |
| Blood urea nitrogen | 2.5 |
| Chronic heart failure | 2.5 |
| COPD | 2.5 |
| Créatinine(umol/L) | 2.5 |
| Face trauma | 2.5 |

COPD: chronic obstructive pulmonary disease

**Supplementary table 6**: risk factors associated with *S. aureus* VAP in the brain-injured population

| | Total population N = 407 | VAP not involving *S. aureus* N = 222 | VAP involving *S. aureus* N = 185 | OR [IC95%] | p-value |
|---|---|---|---|---|---|
| | **n (%) ou m ean ± sd ; P50 [P25 ; P75]** | **n (%) ou mean ± sd ; P50 [P25 ; P75]** | **n (%) ou mean ± sd ; P50 [P25 ; P75]** | | |
| **Sex** | | | | 1.00 | |
| Male | 134 (67.7) | 52 (68.4) | 82 (67.2) | 1.06 [0.57 ; 1.97] | |
| Female | 64 (32.3) | 24 (31.6) | 40 (32.8) | 0.98 [0.96 ; 0.99] | **0.0071** |
| **Age (years)** | 52 [36 ; 63] | 56 [45 ; 63] | 51 [34 ; 59] (18 - 81) | 1.04 [0.98 ; 1.1] | 0.21 |
| **BMI (kg/m2)** | 24.00 [21.17 ; 27.80] | 23.36 [20.70 ; 27.00] | 24.52 [22.10 ; 28.10] (13.50 - 45.80) | 0.79 [0.42 ; 1.47] | 0.45 |
| **Smoking habits** | 66 (37.7) | 28 (41.2) | 38 (35.5) | 0.69 [0.35 ; 1.39] | 0.30 |
| **Alcohol abuse** | 44 (25.1) | 20 (29.4) | 24 (22.4) | 0.2 [0.01 ; 1.59] | **0.13** |
| **Heart failure** | 4 (2.0) | 3 (4.0) | 1 (0.8) | - | **0.049** |
| **Arteritis** | 2 (1.0) | 2 (2.7) | 0 (0.0) | | 0.37 |
| **Diabetes mellitus** | | | | 1.00 | |
| No | 175 (89.3) | 65 (86.7) | 110 (90.9) | 1.00 | |

| | Total population N = 407 | VAP not involving *S. aureus* N = 222 | VAP involving *S. aureus* N = 185 | OR [IC95%] | p-value |
|---|---|---|---|---|---|
| | n (%) ou m ean ± sd ; P50 [P25 ; P75] | n (%) ou mean ± sd ; P50 [P25 ; P75] | n (%) ou mean ± sd ; P50 [P25 ; P75] | | |
| Uncomplicated diabetes | 13 (6.6) | 5 (6.7) | 8 (6.6) | 0.95 [0.3 ; 3.24] | |
| Complicated diabetes | 8 (4.1) | 5 (6.7) | 3 (2.5) | 0.35 [0.07 ; 1.49] | |
| **Hypertension** | 29 (14.8) | 12 (16.0) | 17 (14.0) | 0.86 [0.39 ; 1.95] | 0.71 |
| **Atrial fibrillation** | 3 (1.5) | 2 (2.7) | 1 (0.8) | 0.3 [0.01 ; 3.23] | 0.32 |
| **COPD** | 5 (2.6) | 4 (5.3) | 1 (0.8) | 0.15 [0.01 ; 1.02] | **0.05** |
| **Chronic respiratory failure** | 1 (0.5) | 1 (1.3) | 0 (0.0) | - | **0.16** |
| **Long-term oxygen** | 0 (0.0) | 0 (0.0) | 0 (0.0) | - | - |
| **Cirrhosis** | 1 (0.5) | 1 (1.3) | 0 (0.0) | - | **0.16** |
| **Renal failure** | | | | | 0.35 |
| None | 184 (93.9) | 68 (90.7) | 116 (95.9) | 1.00 | |
| Chronic renal failure without dialysis requirement | 7 (3.6) | 4 (5.3) | 3 (2.5) | 0.44 [0.08 ; 2.05] | |
| Chronic dialysis | 6 [4 ; 8] | 7 [3 ; 9] | 6 [4 ; 8] | 0.93 [0.85 ; 1.02] | **0.12** |

| | Total population N = 407 | VAP not involving *S. aureus* N = 222 | VAP involving *S. aureus* N = 185 | OR [IC95%] | p-value |
|---|---|---|---|---|---|
| | n (%) ou m ean ± sd ; P50 [P25 ; P75] | n (%) ou mean ± sd ; P50 [P25 ; P75] | n (%) ou mean ± sd ; P50 [P25 ; P75] | | |
| **Glasgow coma score** | 8 (4.1) | 3 (4.0) | 5 (4.1) | 1.03 [0.25 ; 5.17] | 0.96 |
| **SAPS II** | 5 (2.5) | 1 (1.3) | 4 (3.3) | 2.54 [0.37 ; 50.24] | 0.37 |
| **Immunodepression** | 7 (3.6) | 4 (5.3) | 3 (2.5) | 0.44 [0.08 ; 2.05] | |
| **COVID-19** | 6 [4 ; 8] | 7 [3 ; 9] | 6 [4 ; 8] (3 - 15) | 0.93 [0.85 ; 1.02] | **0.12** |
| **Chest trauma** | 28 (14.1) | 28 (14.1) | 20 (16.4) | 1.67 [0.72 ; 4.22] | 0.24 |
| **Facial trauma** | 45 (22.7) | 45 (22.7) | 27 (22.1) | 0.92 [0.47 ; 1.83] | 0.80 |
| **Previous antibiotic therapy : Beta-lactamine** | 34 (17.3) | 16 (21.3) | 18 (14.9) | 0.64 [0.31 ; 1.37] | 0.25 |
| **Glycopeptide** | | | | | - |
| No | 196 (100.0) | 75 (100.0) | 121 (100.0) | - | |
| **Aminoside** | | | | | - |
| No | 196 (100.0) | 75 (100.0) | 121 (100.0) | - | |
| **Intracranial pressure monitoring** | | | | | **0.14** |
| None | 21 (10.8) | 11 (14.9) | 10 (8.3) | 1.00 | |

| | Total population N = 407 | | VAP not involving *S. aureus* N = 222 | | VAP involving *S. aureus* N = 185 | OR [IC95%] | p-value |
|---|---|---|---|---|---|---|---|
| | **n (%) ou m ean ± sd ; P50 [P25 ; P75]** | | **n (%) ou mean ± sd ; P50 [P25 ; P75]** | | **n (%) ou mean ± sd ; P50 [P25 ; P75]** | | |
| Parenchymal monitoring | 95 (49.0) | | 39 (52.7) | | 56 (46.7) | 1.58 [0.61 ; 4.14] | |
| External ventricular drain | 78 (40.2) | | 24 (32.4) | | 54 (45.0) | 2.47 [0.93 ; 6.72] | |
| **Hypothermia at admission (< 35°C)** | 12 (6.2) | | 7 (9.3) | | 5 (4.2) | 0.43 [0.12 ; 1.39] | **0.16** |
| **Subarachnoid hemorrhage** | 53 (26.8) | | 53 (26.8) | | 37 (30.3) | 1.63 [0.84 ; 3.27] | **0.15** |
| **Stroke** | 20 (10.1) | | 20 (10.1) | | 10 (8.2) | 0.59 [0.23 ; 1.51] | 0.27 |
| **Haemorrhagic stroke** | 42 (21.2) | | 42 (21.2) | | 27 (22.1) | 1.16 [0.58 ; 2.39] | 0.69 |
| **Mean arterial pressure at admission (mmHg)** | 93 [80 ; 107] | | 93 [80 ; 107] | | 92 [80 ; 103] | 0.98 [0.97 ; 1] | **0.0117** |
| **Feeding tube** | | | | | | | - |
| None | 2 (1.6) | | 1 (2.0) | | 1 (1.3) | - | |
| Oro-gastric tube | 65 (50.4) | | 24 (49.0) | | 41 (51.3) | - | |
| Naso-gastric tube | 62 (48.1) | | 24 (49.0) | | 38 (47.5) | - | |
| **Delay from admission to VAP (days)** | 6 [4 ; 8] | 76 | 6 [4 ; 8] | 122 | 6 [4 ; 8] | 0.98 [0.92 ; 1.06] | 0.66 |

| | Total population N = 407 | VAP not involving *S. aureus* N = 222 | VAP involving *S. aureus* N = 185 | OR [IC95%] | p-value |
|---|---|---|---|---|---|
| | n (%) ou m ean ± sd ; P50 [P25 ; P75] | n (%) ou mean ± sd ; P50 [P25 ; P75] | n (%) ou mean ± sd ; P50 [P25 ; P75] | | |
| **Albumin (g/L)** | 36.1 ± 5.9 | 11 35.7 ± 7.5 | 18 36.4 ± 4.9 | - | - |
| **Leucocytes (G/L)** | 13.20 [10.40 ; 17.70] | 13.15 [10.25 ; 17.60] | 13.20 [10.50 ; 17.70] | 1.01 [0.96 ; 1.06] | 0.68 |
| **CPK (UI/L)** | 336 [152 ; 757] | 331 [120 ; 891] | 341 [156 ; 700] | - | - |
| **Bilirubin (Umol/L)** | 9 [7 ; 13] | 9 [6 ; 13] | 10 [7 ; 13] | - | - |
| **Glycemia (mmol/L)** | 7.63 [6.50 ; 9.70] | 7.58 [6.30 ; 9.20] | 7.70 [6.60 ; 9.90] | - | - |
| **Natremia (mmol/L)** | 138 [136 ; 141] | 139 [135 ; 142] | 138 [136 ; 140] | 1 [0.94 ; 1.05] | 0.86 |
| **Creatinin (µmol/L)** | 72 [57 ; 94] | 71 [57 ; 99] | 74 [57 ; 92] | 1 [1 ; 1] | **0.13** |
| **Blood urea nitrogen (mmol/L)** | 4.75 [3.70 ; 6.16] | 5.05 [3.85 ; 7.00] | 4.60 [3.65 ; 5.90] | 0.91 [0.83 ; 0.98] | **0.0090** |
| **Proton pump inhibitors** | 32 (50.0) | 13 (48.1) | 19 (51.4) | - | - |
| **Systemic corticosteroid** | 53 (57.6) | 15 (45.5) | 38 (64.4) | - | - |
| **Insulin** | 82 (100.0) | 34 (100.0) | 48 (100.0) | - | - |
| **Per-surgery antibioprophylaxis** | 63 (34.8) | 28 (41.2) | 35 (31.0) | 0.64 [0.34 ; 1.2] | **0.16** |
| **Pentothal** | 33 (16.7) | 7 (9.2) | 26 (21.3) | .67 [1.15 ; 6.99] | **0.0214** |
| **Midazolam** | 146 (75.3) | 48 (65.8) | 98 (81.0) | 2.22 [1.14 ; 4.33] | **0.0184** |

| | Total population N = 407 | VAP not involving *S. aureus* N = 222 | VAP involving *S. aureus* N = 185 | OR [IC95%] | p-value |
|---|---|---|---|---|---|
| | n (%) ou m ean ± sd ; P50 [P25 ; P75] | n (%) ou mean ± sd ; P50 [P25 ; P75] | n (%) ou mean ± sd ; P50 [P25 ; P75] | | |
| **Blood transfusion** | | | | | - |
| none | 69 (52.7) | 29 (56.9) | 40 (50.0) | - | |
| <= 3 | 45 (34.4) | 15 (29.4) | 30 (37.5) | - | |
| > 3 | 17 (13.0) | 7 (13.7) | 10 (12.5) | - | |
| **Delay from admission to tracheal intubation** | 1 [1 ; 1] | 1 [1 ; 1] | 1 [1 ; 1] | 0.99 [0.97 ; 1.02] | 0.68 |
| **Length of invasive mechanical ventilation (days)** | 21 [13 ; 31] | 21 [13 ; 31] | 20 [14 ; 32] | 1 [0.99 ; 1.02] | 0.84 |
| **Prone position** | 24 (12.1) | 8 (10.5) | 16 (13.1) | 1.28 [0.53 ; 3.31] | 0.58 |
| **Normothermia** | 59 (29.9) | 17 (22.7) | 42 (34.4) | 1.79 [0.94 ; 3.52] | **0.08** |
| **Chest surgery** | 9 (4.5) | 4 (5.3) | 5 (4.1) | 0.77 [0.2 ; 3.2] | 0.70 |
| **Neurochirurgy** | 147 (74.2) | 56 (73.7) | 91 (74.6) | 1.05 [0.54 ; 2.01] | 0.89 |
| **Orthopedic surgery** | 17 (8.6) | 9 (11.8) | 8 (6.6) | 0.52 [0.19 ; 1.43] | 0.20 |

**Supplementary table 7**: backward multivariate analysis in the non-brain-injured population: percentage of times the variable is selected in the model

| **Variable** | percentage of times the variable is selected in the model (40 réplications) **(%)** |
|---|---|
| Antibioprophylaxis | 100 |
| Age (years) | 100 |
| Arteritis | 100 |
| SAPS II | 100 |
| Thoracic surgery | 100 |
| Neurosurgery | 100 |
| Length of invasive mechanical ventilation | 57.5 |
| Creatinin (µmol/l) | 32.5 |
| Initial mean arterial pressure | 17.5 |

**Supplementary table 8**: risk factors associated with *S. aureus* VAP in the non-brain-injured population

| | Total population n = 407 | VAP not involving *S. aureus* n = 222 | VAP involving *S. aureus* n = 185 | OR [IC95%] | p-value |
|---|---|---|---|---|---|
| | **n (%) or mean ± sd; P50 [P25; P75]** | **n (%) or mean ± sd; P50 [P25; P75]** | **n (%) or mean ± sd; P50 [P25; P75]** | | |
| **Sex** | | | | 1.19 [0.62 ; 2.25] | 0.59 |
| Male | 148 (70.8) | 105 (71.9) | 43 (68.3) | | |
| Female | 61 (29.2) | 41 (28.1) | 20 (31.7) | | |
| **Age (years)** | 64 [55 ; 73] (19 - 95) | 66 [58 ; 75] (19 - 95) | 59 [51 ; 66] (20 - 79) | 0.95 [0.93 ; 0.97] | **< 0.0001** |
| **BMI (kg/m2)** | 27.15 [23.40 ; 31.60] | 27.20 [23.40 ; 31.60] (14.10 - 50.00) | 26.80 [22.60 ; 31.30] (17.40 - 46.20) | 0.98 [0.94 ; 1.03] | 0.50 |
| **Smoking habits** | 53 (27.5) | 38 (27.7) | 15 (26.8) | 0.95 [0.46 ; 1.89] | 0.89 |
| **Alcohol abuse** | 38 (19.7) | 27 (19.7) | 11 (19.6) | 1 [0.44 ; 2.13] | 0.99 |
| **Heart failure** | 17 (8.1) | 14 (9.6) | 3 (4.8) | 0.47 [0.11 ; 1.51] | 0.22 |
| **Arteritis** | 15 (7.2) | 15 (10.3) | 0 (0.0) | - | **0.0008** |
| **Diabetes mellitus** | | | | | 0.47 |
| No | 165 (78.9) | 112 (76.7) | 53 (84.1) | 1.00 | |

| | Total population n = 407 | VAP not involving *S. aureus* n = 222 | VAP involving *S. aureus* n = 185 | OR [IC95%] | p-value |
|---|---|---|---|---|---|
| | n (%) or mean ± sd; P50 [P25; P75] | n (%) or mean ± sd; P50 [P25; P75] | n (%) or mean ± sd; P50 [P25; P75] | | |
| Uncomplicated diabetes | 27 (12.9) | 21 (14.4) | 6 (9.5) | 0.6 [0.21; 1.5] | |
| Complicated diabetes | 17 (8.1) | 13 (8.9) | 4 (6.3) | 0.65 [0.18; 1.94] | |
| **Hypertension** | 58 (27.8) | 40 (27.4) | 18 (28.6) | 1.06 [0.54; 2.03] | 0.86 |
| **Atrial fibrillation** | 11 (5.3) | 8 (5.5) | 3 (4.8) | 0.86 [0.18; 3.1] | 0.32 |
| **COPD** | 12 (5.7) | 10 (6.8) | 2 (3.2) | 0.45 [0.07; 1.76] | **0.05** |
| **Chronic respiratory failure** | 0 (0.0) | 0 (0.0) | 0 (0.0) | - | **0.16** |
| **Long-term oxygen** | 0 (0.0) | 0 (0.0) | 0 (0.0) | - | - |
| **Cirrhosis** | 2 (1.0) | 2 (1.4) | 0 (0.0) | - | **0.16** |
| **Renal failure** | | | | | 0.35 |
| None | 189 (90.4) | 128 (87.7) | 61 (96.8) | 1.00 | |
| Chronic renal failure without dialysis requirement | 19 (9.1) | 17 (11.6) | 2 (3.2) | 0.25 [0.04; 0.9] | |
| Chronic dialysis | 1 (0.5) | 1 (0.7) | 0 (0.0) | - | |
| **Glasgow coma score** | 15 [15; 15] (3 - 15) | 15 [15; 15] (3 - 15) | 15 [14; 15] (3 - 15) | 1.02 [0.91; 1.16] | 0.75 |
| **SAPS II** | 44 [36; 65] (15 - 103) | 50 [37; 68] (15 - 103) | 39 [32; 58] (18 - 94) | 0.98 [0.96; 0.99] | **0.0082** |

| | Total population<br>n = 407 | VAP not involving *S. aureus*<br>n = 222 | VAP involving *S. aureus*<br>n = 185 | OR [IC95%] | p-value |
|---|---|---|---|---|---|
| | n (%) or mean ± sd; P50 [P25; P75] | n (%) or mean ± sd; P50 [P25; P75] | n (%) or mean ± sd; P50 [P25; P75] | | |
| **Immunodepression** | 36 (17.2) | 25 (17.1) | 11 (17.5) | 1.02 [0.45; 2.19] | 0.95 |
| **COVID-19** | 70 (33.5) | 39 (26.7) | 31 (49.2) | 2.66 [1.44; 4.94] | **0.0018** |
| **Chest trauma** | 24 (11.5) | 16 (11.0) | 8 (12.7) | 1.18 [0.46; 2.85] | 0.72 |
| **Facial trauma** | 4 (1.9) | 1 (0.7) | 3 (4.8) | 7.25 [0.91; 148.17] | **0.06** |
| **Previous antibiotic therapy : Beta-lactamine** | 117 (56.0) | 80 (54.8) | 37 (58.7) | 1.17 [0.65; 2.15] | 0.60 |
| **Glycopeptide** | | | | | - |
| No | | 146 (100.0) | - | | |
| **Aminoside** | | | | | - |
| No | 209 (100.0) | 146 (100.0) | 63 (100.0) | - | |
| **Hypothermia at admission (< 35°C)** | 7 (3.5) | 6 (4.2) | 1 (1.8) | 0.41 [0.02; 2.46] | 0.37 |
| **Sepsis / septic shokc** | 10 (4.8) | 8 (5.5) | 2 (3.2) | 0.57 [0.08; 2.34] | 0.46 |
| **Mean arterial pressure at admission (mmHg)** | 85 [72 ; 100] | 81 [72 ; 98] | 95 [80 ; 102] | 1.02 [1.01 ; 1.04] | **0.0074** |

| | Total population n = 407 | VAP not involving *S. aureus* n = 222 | VAP involving *S. aureus* n = 185 | OR [IC95%] | p-value |
|---|---|---|---|---|---|
| | n (%) or mean ± sd; P50 [P25; P75] | n (%) or mean ± sd; P50 [P25; P75] | n (%) or mean ± sd; P50 [P25; P75] | | |
| **Feeding tube** | | | | | - |
| None | 2 (1.7) | 2 (2.4) | 0 (0.0) | - | |
| Oro-gastric tube | 72 (61.0) | 50 (61.0) | 22 (61.1) | - | |
| Naso-gastric tube | 44 (37.3) | 30 (36.6) | 14 (38.9) | - | |
| **Delay from admission to VAP (days)** | 6 [4 ; 9] | 6 [4 ; 9] | 6 [5 ; 10] | 1.01 [0.97; 1.05] | 0.63 |
| **Albumin (g/L)** | 28.8 ± 6.2 | 28.9 ± 6.7 | 28.4 ± 4.6 | - | - |
| **Leucocytes (G/L)** | 12.00 [8.20 ; 17.41] (0.79 - 322.46) | 12.45 [8.05; 17.46] (0.79 - 70.17) | 11.80 [8.65; 17.10] (2.28 - 322.46) | 1.01 [1; 1.04] | 0.20 |
| **CPK (UI/L)** | 350 [125 ; 981] (39 - 138,384) | 299 [113; 663] (39 - 138,384) | 844 [173; 3,527] (88 - 50,338) | - | - |
| **Bilirubin (Umol/L)** | 12 [7; 18] (3 - 84) | 12 [7; 18] (3 - 84) | 13 [7; 18] (4 - 23) | - | - |
| **Glycemia (mmol/L)** | 8.10 [6.60; 11.00] (2.50 - 32.69) | 8.10 [6.60; 11.00] (2.50 - 27.00) | 7.86 [6.70; 11.45] (3.30 - 32.69) | - | - |
| **Natremia (mmol/L)** | 137 [134 ; 139] (115 - 152) | 137 [133 ; 139] (115 - 152) | 137 [134 ; 139] (120 - 147) | 1.01 [0.96; 1.08] | 0.67 |

| | Total population n = 407 | VAP not involving *S. aureus* n = 222 | VAP involving *S. aureus* n = 185 | OR [IC95%] | p-value |
|---|---|---|---|---|---|
| | **n (%) or mean ± sd; P50 [P25; P75]** | **n (%) or mean ± sd; P50 [P25; P75]** | **n (%) or mean ± sd; P50 [P25; P75]** | | |
| **Creatinin (µmol/L)** | 96 [70 ; 141] (29 - 520) | 97 [75 ; 142] (44 - 520) | 94 [67 ; 135] (29 - 410) | 1 [0.99; 1] | **0.19** |
| **Blood urea nitrogen (mmol/L)** | 8.10 [5.40 ; 12.50] (3.00 - 66.50) | 8.60 [6.00 ; 12.70] (3.00 - 44.00) | 6.85 [5.00 ; 10.70] (3.30 - 66.50) | 0.98 [0.94; 1.02] | 0.37 |
| **Proton pump inhibitors** | 65 (91.5) | 45 (90.0) | 20 (95.2) | - | - |
| **Systemic corticosteroid** | 111 (88.1) | 76 (88.4) | 35 (87.5) | - | - |
| **Insulin** | 74 (98.7) | 59 (100.0) | 15 (93.8) | - | - |
| **Midazolam** | 73 (35.4) | 49 (33.8) | 24 (39.3) | 1.27 [0.68; 2.35] | 0.45 |
| **Dialysis during ICU stay** | 93 (44.5) | 68 (46.6) | 25 (39.7) | 0.75 [0.41; 1.37] | 0.36 |
| **Blood transfusion** | | | | | |
| none | 60 (33.9) | 35 (26.1) | 25 (58.1) | - | |
| <= 3 | 44 (24.9) | 35 (26.1) | 9 (20.9) | - | |
| > 3 | 73 (41.2) | 64 (47.8) | 9 (20.9) | - | |
| **Delay from admission to tracheal intubation** | 1 [1 ; 24] | 1 [1 ; 2] | 1 [1 ; 24] | 1 [1; 1.01] | **0.07** |
| **Length of invasive mechanical ventilation (days)** | 20 [13 ; 33] (2 - 142) | 19 [11 ; 31] | 23 [16 ; 37] | 1.01 [1; 1.03] | **0.07** |

| | Total population n = 407 | VAP not involving *S. aureus* n = 222 | VAP involving *S. aureus* n = 185 | OR [IC95%] | p-value |
|---|---|---|---|---|---|
| | **n (%) or mean ± sd; P50 [P25; P75]** | **n (%) or mean ± sd; P50 [P25; P75]** | **n (%) or mean ± sd; P50 [P25; P75]** | | |
| **Prone position** | 111 (53.1) | 73 (50.0) | 38 (60.3) | 1.52 [0.84; 2.79] | **0.17** |
| **Normothermia** | 0 (0.0) | 0 (0.0) | 0 (0.0) | - | - |
| **Chest surgery** | 61 (29.2) | 57 (39.0) | 4 (6.3) | 0.11 [0.03; 0.28] | **< 0.0001** |
| **Neurochirurgy** | 1 (0.5) | 0 (0.0) | 1 (1.6) | - | **0.12** |
| **Orthopedic surgery** | 19 (9.1) | 11 (7.5) | 8 (12.7) | 1.79 [0.66; 4.65] | 0.25 |

**Supplementary figure 1**: Forest plot of risk factors of involvement of *Staphylococcus aureus* in VAP in the whole population: multivariate analysis

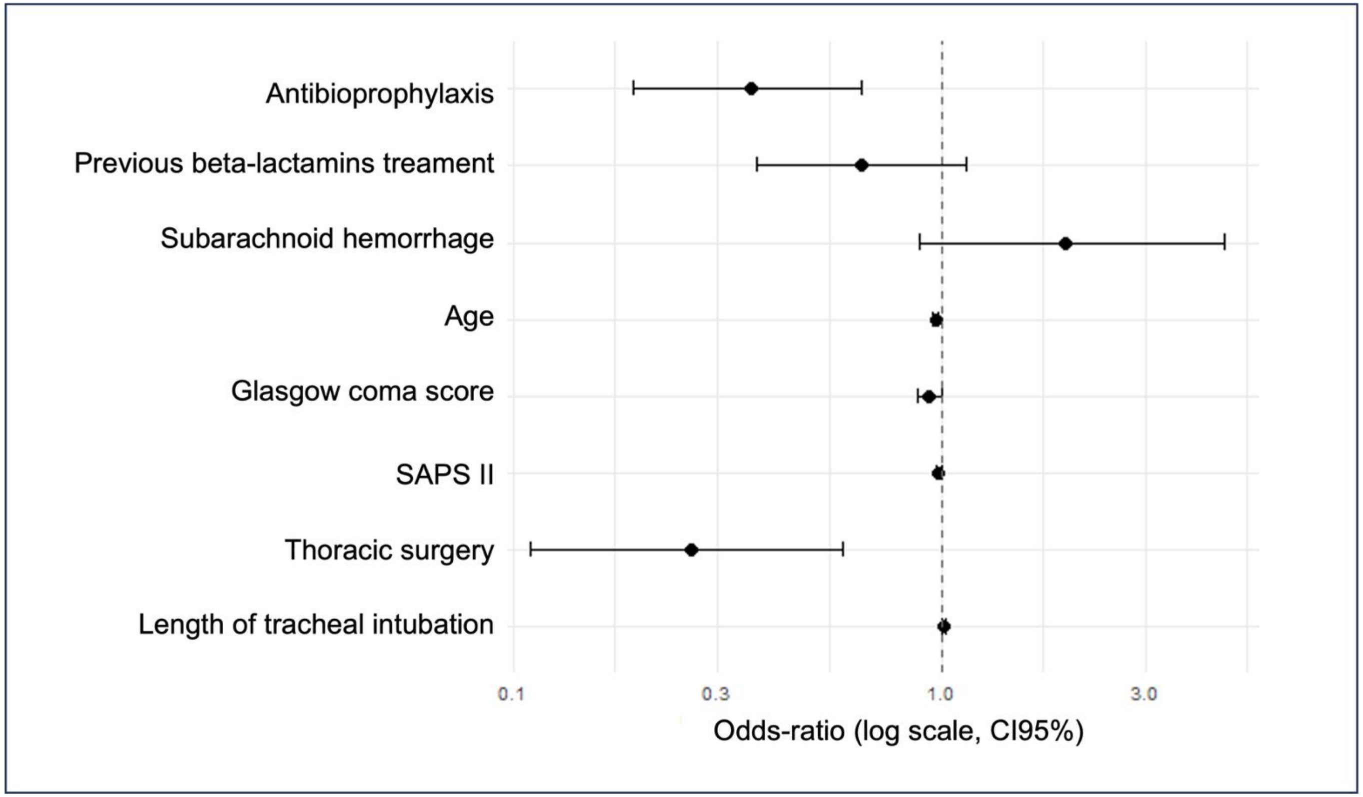
